# Amount and intensity of physical activity and risk of incident cancer in the UK Biobank

**DOI:** 10.1101/2023.12.04.23299386

**Authors:** Alaina H. Shreves, Scott R. Small, Rosemary Walmsley, Shing Chan, Pedro F. Saint-Maurice, Steven C. Moore, Keren Papier, Kezia Gaitskell, Ruth C. Travis, Charles E. Matthews, Aiden Doherty

## Abstract

**Importance:** The influence of total daily and light intensity activity on cancer risk remains unclear, as most existing knowledge is drawn from studies relying on self-reported leisure-time activities of moderate-vigorous intensity.

**Objective:** To investigate associations between total daily activity, including step counts, and activity intensity on incident cancer risk.

**Design, Setting, and Participants:** Prospective analysis of cancer-free UK Biobank participants who wore accelerometers for 7-days (between 2013-2015), followed for cancer incidence through national registries (mean follow-up 5.8 years (SD=1.3)).

**Exposures:** Time-series machine learning models derived daily total activity (average acceleration), behaviour time, step counts, and peak 30-minute cadence from wrist-based accelerometer data.

**Main Outcomes and Measures:** A composite cancer outcome of 13 cancers previously associated with low physical activity (bladder, breast, colon, endometrial, oesophageal adenocarcinoma, gastric cardia, head and neck, kidney, liver, lung, myeloid leukaemia, myeloma, and rectum) based on previous studies of self-reported activity. Cox proportional hazards regression models estimated hazard ratios (HR) and 95% confidence intervals (CI), adjusted for age, sex, ethnicity, smoking, alcohol, education, Townsend Deprivation Index, and reproductive factors. Associations of reducing sedentary time in favour of increased light and moderate-vigorous activity were examined using compositional data analyses.

**Results:** Among 86 556 participants (mean age 62.0 years (SD=7.9) at accelerometer assessment), 2 669 cancers occurred. Higher total physical activity was associated with a lower overall cancer risk (HR_1SD_=0.85, [95%CI 0.81-0.89]). On average, reallocating one hour/day from sedentary behaviour to moderate-vigorous physical activity was associated with a lower risk (HR=0.92, [0.89-0.95]), as was reallocating one hour/day to light-intensity physical activity (HR=0.94, [0.92-0.96]). Compared to individuals taking 5 000 daily steps, those who took 9 000 steps had an 18% lower risk of physical-activity-related cancer (HR=0.82, [0.74-0.90]). We found no significant association with peak 30-minute cadence after adjusting for total steps.

**Conclusion and Relevance:** Higher total daily physical activity and less sedentary time, in favour of both light and moderate-vigorous intensity activity, were associated with a lower risk of certain cancers. For less active adults, increasing step counts by 4 000 daily steps may be a practical public health intervention for lowering the risk of some cancers.

**KEY POINTS:** *Question:* What insights can we gain about the relationships between total daily activity, step counts, and activity intensity on cancer risk using accelerometer data?

*Findings:* In an analysis of 86 556 individuals from the UK Biobank who provided a week of accelerometer-based activity data, higher levels of total physical activity, reducing sedentary time in favour of light or moderate-vigorous intensity activities, and higher daily step counts were associated with a lower risk of certain cancers.

*Meaning:* For less active adults, increasing activity time and accumulating more daily steps may be practical interventions for lowering the risk of some cancers.

## INTRODUCTION

Epidemiological data indicate that over half of all new cancers in high-income countries could be avoided by modifying lifestyle factors, including addressing physical inactivity.^1,2^ However, quantifying the dose-dependent benefits of specific physical activity (PA) behaviours for cancer prevention remains challenging. Many existing studies rely on self-report questionnaires, which emphasize time spent in leisure-time activities of a moderate-vigorous intensity and may have recall errors and reporting biases.^3,4^ Time use data studies suggest that most individuals spend a significant amount of time devoted to work and household activities, with the majority of physically active time obtained from light-intensity physical activity (LIPA).^5–7^ Global adherence to PA recommendations, which primarily emphasize moderate-vigorous physical activity (MVPA), is generally poor.^8^ Consequently, there is a growing interest in understanding whether more accurate measures of MVPA show protective effects for cancer risk and if interventions to increase LIPA could be a valuable disease prevention strategy.

Wearable accelerometer devices provide objective measures of all daily activities, including sedentary and physically active time accumulated at home, work, during transportation, and in leisure-time.^9^ Accelerometer data can also be used to investigate how reallocating time from sedentary behaviour to either LIPA or MVPA could influence disease risk. Existing accelerometer-measured activity studies in cancer have primarily focused on cancer mortality and,^10–14^ to a limited extent, breast cancer risk.^15,16^

To address these gaps and challenges, we aimed to understand the relationship between total daily activity and intensity, step counts, and incident cancer risk using a composite outcome of 13 cancer sites previously associated with low PA in studies of self-reported leisure time activity.^17^ As a secondary analysis, we assessed the risks for all 13 PA-related cancers individually and, as case numbers allowed, for other site-specific cancers. We also conducted compositional data analyses to explore the impact of reallocating time from sedentary behaviour to LIPA and MVPA. Finally, for potential translation to the clinical and public health settings, we examined the dose-response relationship between step count, stepping intensity, and incident cancer. Step counts are an easily understandable metric among the general population and are reported by many consumer wearable devices and fitness trackers.^18^

## METHODS

### Study population

The UK Biobank is a prospective study that enrolled 502 536 adults in England, Scotland, and Wales between 2006-2010.^19,20^ At baseline, participants completed a questionnaire, provided biological data, and consented for linkage to electronic medical records. From June 2013 to December 2015, participants with valid emails were invited to wear an Axivity AX3 wrist-worn accelerometer for 7 days.^7^

### Accelerometer data processing

Accelerometer data were processed using methods described by Doherty (“accelerometer”, v7.1.0).^21^ Total physical activity (PA) was calculated as the mean vector magnitude per epoch to derive an overall mean per day acceleration in milligravity (m*g*) units. This metric reflects the activity duration and intensity and has been validated against doubly labelled water.^21^

Proportions of time spent across sleep, sedentary behaviour (SB), light-intensity physical activity (LIPA), and moderate-vigorous physical activity (MVPA) per day, were calculated using random forest and hidden Markov model machine-learning methods.^7^ Missing time due to non-wear was imputed by averaging the behaviour in the corresponding times across all valid days.^7^

Step counts were calculated using a hybrid self-supervised learning model trained on ground truth free-living stepping data (“stepcount”, v3.1.1).^22^ Daily step count was reported as the median number of daily steps during the seven-day measurement period. Peak 30-minute cadence was calculated as the mean of the 30 highest daily cadence values, averaged across all days.^23^ Further processing details are described in eTable 1.

### Outcome ascertainment

The main outcome was a composite cancer outcome of 13 sites previously found to be associated with low PA (bladder, breast, colon, endometrial, oesophageal adenocarcinoma, gastric cardia, head and neck, kidney, liver, lung, myeloid leukaemia, myeloma, and rectal).^17^ Cancers were obtained by the UK Biobank through the National Health Service (NHS) Digital for participants from England and Wales and the NHS Central Register for participants from Scotland (details in eTable 2).^24^ In secondary analyses, we assessed the risks for cancers not previously related to PA with at least 100 cases (eTable 3).

### Analytic sample

Raw accelerometer data from 103 614 participants were processed, excluding study withdrawals. We further excluded participants with device calibration or data reading errors (>1% of values outside +/-8*g* range), inadequate wear time (<72 hours), unreasonably high average acceleration (>100 m*g*), and lacking steps data.{Citation} Individuals with cancer (excluding C44: non-melanoma skin cancer) before accelerometer wear and missing healthcare linkages or covariate data, were also excluded. The final analysis included 86 556 participants (eFigure 1).

**Figure 1.**
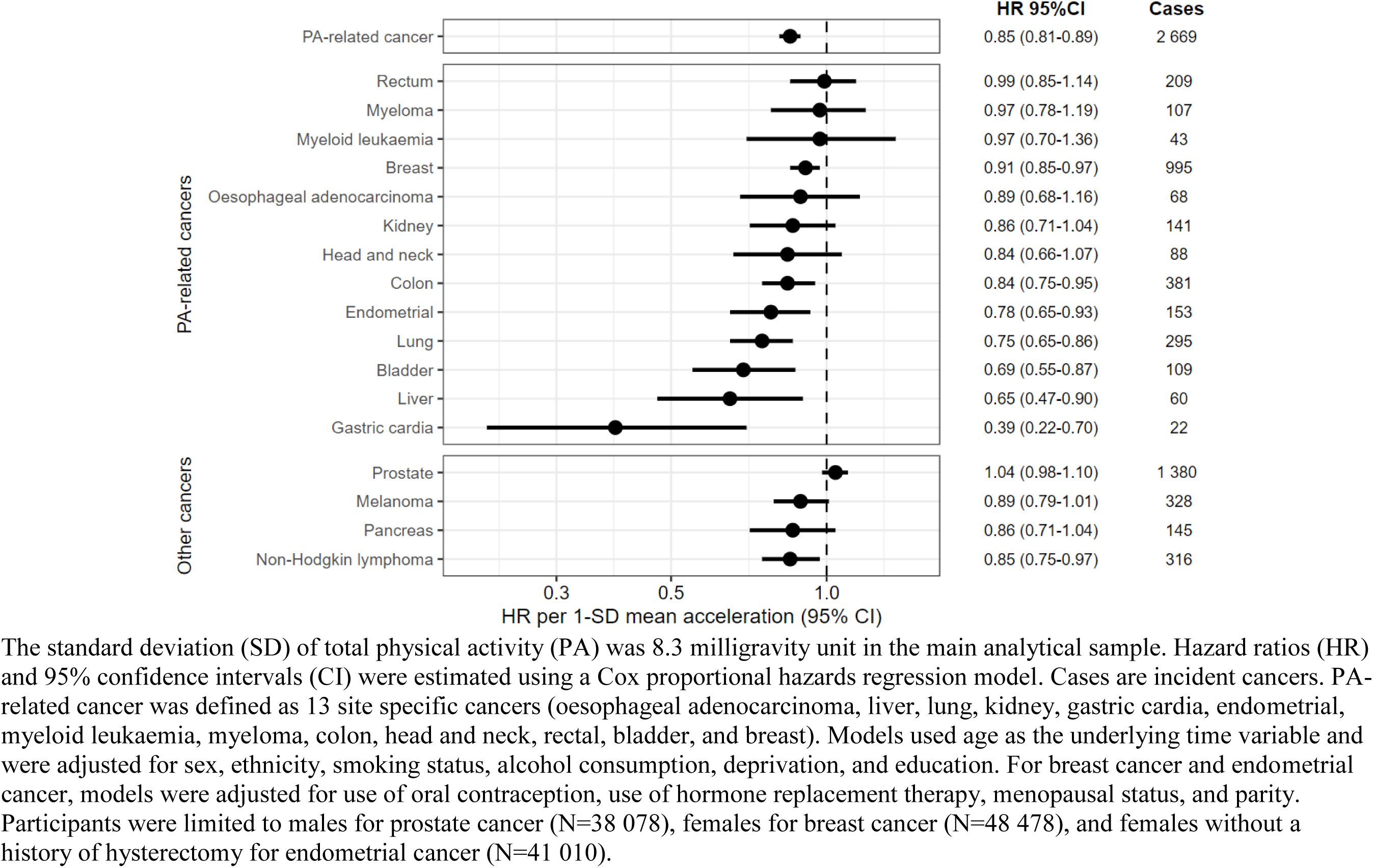
Association of mean accelerometer-measured physical activity with risk of incident cancers in 86 556 UK Biobank participants.

### Statistical analysis

Cox proportional hazards regression models estimated adjusted Hazard ratios (HRs) with 95% confidence intervals for a one standard deviation (SD) increase in total activity and incident cancer. We also assessed risks across quintiles of physical activity.

Attained age was the underlying time scale. Individuals who were cancer-free during follow-up were censored at their date of death or at the end of the follow-up period (31 December 2020, England; 31 December 2016, Wales; 30 November 2021 Scotland).

Multivariable models adjusted for sex (male, female), ethnicity (White, non-White), smoking status (never, previous, current unknown cigarettes/day, current <15 cigarettes/day, current ≥15 cigarettes/day), alcohol consumption (never, <3 times per week, ≥3 times per week), education (school leaver, further education, higher education), and deprivation based on the Townsend Deprivation Index (TDI) (quintiles ranging from least deprived to most deprived, based on the 2011 UK census).^25^ Female-specific models were further adjusted for oral contraception ever use (no, yes, missing), hormone replacement therapy ever use (no, yes, missing), menopausal status (no, yes, missing/unknown), and parity (0, 1-2, 3+, missing). Covariate data were provided by participants at the UK Biobank baseline assessment and were chosen a priori based on existing studies (Details in eTable 1). No violations of the proportional hazards assumption were observed for the exposures using Schoenfeld residuals.

Compositional data analyses, using the "epicoda" package, modelled associations between the relative time spent in SB, LIPA, MVPA, and sleep and cancer risk.^7^ First, we calculated the estimated HR associated with reallocating time to one behaviour from all other proportionally (e.g., reallocating one hour/day to LIPA from all other behaviours proportionally). ^7^ Second, we estimated HRs for specific pairwise reallocations of time between behaviours (e.g., reallocating one hour/day from SB to LIPA, holding sleep and MVPA constant). For all compositional analyses, the estimated hazard ratios were relative to the mean behaviour composition among included participants for a hypothetical average participant.

Finally, we used restricted cubic splines to assess the relationship between step count and cancer incidence using a reference point at the 10th percentile and knots positioned at the 5th, 50th, and 90th percentiles. We employed *P*-values for linear trends to estimate significance of the overall association. Multivariable models were further adjusted for daily step count.

We conducted several sensitivity analyses, including adjustment for body mass index (<25, 25-30, and 30+ kilograms/meters²) as measured at the assessment centre and dietary factors (fresh fruits and vegetable consumption (<3, 3–4.9, 5–7.9, or 8+ servings/day, missing); red and processed meat consumption (<1, 1–2.9, 3–4.9, or 5+ times/week, missing)). Two subgroup analyses were conducted, one among males and females, and the other among never smokers. To assess the potential influence of reverse causality, we repeated the analyses after excluding the first two years of follow-up. Statistical analyses were performed May-November 2023 using R (v4.2.2; R Foundation for Statistical Computing). Two-sided P values of <0.05 was considered statistically significant.

### Consent

Participants in the UK Biobank provided written informed consent. The study was approved by the National Information Governance Board for Health and Social Care and the National Health Service North West Multicentre Research Ethics Committee (06/MRE08/65). We adhered to the Strengthening the Reporting of Observational Studies in Epidemiology (STROBE) reporting guidelines (eTable 4).^26^

## RESULTS

Among 86 556 participants, the mean age at accelerometer assessment was 62.0 years (SD=7.9). The majority were female (56%), identified as White (97%), and fell within the least deprived quintile of the Townsend Deprivation Index (50%) (Table 1). Throughout a mean follow-up of 5.8 years (SD=1.3; 504 557 person-years), 2 669 physical activity (PA)-related cancers accrued, with breast among females (n=995) being the most common (details in eTable 3).

**Table 1.** Overall physical activity metrics by demographic characteristic in 86 556 UK Biobank participants.

| Characteristic |  | No. (%) | Overall acceleration (mg) | Daily step count Median (IQR) | Peak 30 min cadence (steps/min) Median (IQR) |
| --- | --- | --- | --- | --- | --- |
| **Overall** |  | 86 556 (100) | 27.3 (22.7-32.7) | 9 073 (6 834-11 686) | 91 (81-101) |
| Sex | Female | 48 478 (56) | 27.8 (23.2-33.1) | 9 004 (6 812-11 560) | 92 (82-102) |
|  | Male | 38 078 (44) | 26.7 (22.0-32.3) | 9 152 (6 870-11 878) | 90 (81-100) |
| Age (years) | 40-49 | 7 693 (9) | 30.5 (25.6-36.7) | 9 393 (7 126-12 070) | 95 (85-105) |
|  | 50-59 | 25 659 (30) | 29.1 (24.4-34.8) | 9 204 (6 996-11 913) | 93 (84-103) |
|  | 60-69 | 38 214 (44) | 26.8 (22.3-31.9) | 9 166 (6 922-11 772) | 91 (81-100) |
|  | 70-79 | 14 990 (17) | 24.3 (20.2-28.9) | 8 430 (6 255-10 868) | 87 (77-96) |
| Ethnicity | Non-white | 2 726 (3) | 28.5 (23.7-34.1) | 8 720 (6 508-11 376) | 91 (80-102) |
|  | White | 83 830 (97) | 27.3 (22.6-32.7) | 9 084 (6 845-11 696) | 91 (81-101) |
| Quintiles of Townsend Deprivation Index | Least Deprived | 43 574 (50) | 27.4 (22.8-32.7) | 9 080 (6 922-11 633) | 90 (81-100) |
|  | 2nd Quintile | 19 542 (23) | 27.3 (22.6-32.7) | 9 078 (6 817-11 656) | 91 (81-101) |
|  | 3rd Quintile | 12 151 (14) | 27.4 (22.7-32.9) | 9 164 (6 816-11 862) | 93 (82-103) |
|  | 4th Quintile | 8 381 (10) | 27.0 (22.1-32.7) | 8 970 (6 586-11 802) | 93 (82-103) |
|  | Most Deprived | 2 908 (3) | 26.6 (21.6-32.4) | 8 812 (6 293-11 664) | 94 (81-104) |
| Education qualifications | School leaver | 19 817 (23) | 26.8 (22.0-32.2) | 8 621 (6 428-11 229) | 89 (79-99) |
|  | Further education | 28 993 (33) | 27.3 (22.6-32.7) | 8 883 (6 644-11 543) | 90 (80-100) |
|  | Higher education | 37 746 (44) | 27.6 (23.0-33.0) | 9 434 (7 208-12 009) | 93 (84-103) |
| Smoking status | Never | 49 736 (57) | 27.6 (23.0-33.1) | 9 183 (6 980-11 762) | 92 (82-102) |
|  | Previous smoker | 30 805 (36) | 27.0 (22.4-32.4) | 8 984 (6 720-11 671) | 90 (80-100) |
|  | Current, unknown cig/day | 2 351 (3) | 27.0 (22.3-32.2) | 9 037 (6 766-11 730) | 90 (80-100) |
|  | Current, < 15 cig/day | 899 (1) | 25.6 (21.0-31.1) | 8 160 (5 989-10 690) | 86 (75-97) |
|  | Current, ≥ 15 cig/day | 2 765 (3) | 25.3 (20.5-30.8) | 8 108 (5 808-10 818) | 87 (75-96) |
| Alcohol consumption | Never | 4 859 (6) | 26.4 (21.5-32.1) | 8 530 (6 120-11 313) | 89 (78-100) |
|  | < 3 times/week | 39 335 (45) | 27.1 (22.4-32.5) | 8 762 (6 554-11 379) | 90 (80-100) |
|  | 3+ times/week | 42 362 (49) | 27.6 (23.0-33.0) | 9 411 (7 186-12 000) | 92 (83-102) |
| Body mass index, kg/m<sup>2</sup> | ≤ 24.9 kg/m<sup>2</sup> | 33 234 (38) | 29.4 (24.6-35.2) | 9 754 (7 548-12 401) | 95 (85-105) |
|  | 25-29.9 kg/m<sup>2</sup> | 36 206 (42) | 26.9 (22.5-32.1) | 9 094 (6 916-11 653) | 91 (81-100) |
|  | 30+ kg/m<sup>2</sup> | 17 116 (20) | 24.2 (20.0-29.1) | 7 610 (5 564-10 060) | 85 (74-95) |
| Fruit and vegetable consumption | < 3 servings/day | 3 683 (4) | 25.6 (20.7-30.8) | 8 187 (5 930-10 973) | 88 (78-99) |
|  | 3-4.9 servings/day | 14 167 (16) | 26.7 (22.0-32.0) | 8 781 (6 580-11 342) | 90 (80-100) |
|  | 5-7.9 servings/day | 36 127 (42) | 27.3 (22.7-32.7) | 9 117 (6 918-11 682) | 91 (82-101) |
|  | 8+ servings/day | 31 857 (37) | 27.9 (23.2-33.3) | 9 232 (6 981-11 922) | 92 (82-101) |
|  | Missing | 722 (1) | 26.4 (21.2-32.6) | 8 914 (6 127-11 825) | 90 (79-100) |
| Red and processed meat consumption | Less than 1 time/week | 6 802 (8) | 29.3 (24.3-35.1) | 9 674 (7 256-12 397) | 96 (85-106) |
|  | 1-2.9 times/week | 33 047 (38) | 27.5 (22.9-32.9) | 9 091 (6 906-11 682) | 92 (82-102) |
|  | 3-4.9 times/week | 25 399 (29) | 27.1 (22.5-32.5) | 8 992 (6 768-11 582) | 91 (81-100) |
|  | 5+ times/week | 20 927 (24) | 26.6 (21.9-32.0) | 8 964 (6 676-11 567) | 90 (80-99) |
|  | Missing | 381 (0) | 28.2 (21.5-34.1) | 9 158 (6 190-12 077) | 91 (79-101) |
| Self-reported usual walking pace | Brisk pace | 41 087 (47) | 29.0 (24.3-34.6) | 9 630 (7 422-12 250) | 94 (85-104) |
|  | Steady average pace | 41 381 (48) | 26.2 (21.8-31.2) | 8 737 (6 575-11 294) | 89 (80-99) |
|  | Slow pace | 3 907 (5) | 21.9 (17.6-26.8) | 6 352 (4 150-8 870) | 77 (64-89) |
|  | None of the above | 76 (0) | 23.5 (18.5-29.3) | 5 527 (2 945-9 778) | 78 (54-87) |
|  | Missing | 105 (0) | 18.9 (13.9-23.5) | 3 397 (1 102-5 684) | 56 (26-72) |
| Self-rated overall health | Excellent | 19 332 (22) | 29.2 (24.5-35.0) | 9 765 (7 566-12 386) | 95 (85-104) |
|  | Good | 51 914 (60) | 27.4 (22.8-32.6) | 9 140 (6 946-11 727) | 91 (82-101) |
|  | Fair | 13 079 (15) | 25.0 (20.6-30.2) | 8 100 (5 885-10 650) | 87 (76-97) |
|  | Poor | 2 089 (2) | 22.1 (17.6-27.2) | 6 378 (4 064-8 983) | 78 (64-90) |
|  | Missing | 142 (0) | 23.3 (19.6-28.4) | 7 928 (5 528-10 270) | 85 (75-95) |
| Wear season | Spring | 19 875 (23) | 27.7 (23.0-33.2) | 9 207 (6 946-11 880) | 92 (82-102) |
|  | Summer | 22 946 (27) | 27.9 (23.1-33.4) | 9 514 (7 208-12 186) | 91 (82-101) |
|  | Autumn | 25 518 (29) | 27.2 (22.7-32.6) | 8 966 (6 794-11 522) | 91 (81-101) |
|  | Winter | 18 217 (21) | 26.3 (21.9-31.5) | 8 518 (6 398-11 058) | 90 (80-101) |
| **Females<sup>a</sup>** |  | 48 478 (100) | 27.8 (23.2-33.1) | 9 004 (6 812-11 560) | 92 (82-102) |
| Number of live births | 0 | 10 373 (21) | 27.9 (23.3-33.4) | 9 060 (6 792-11 700) | 94 (83-105) |
|  | 1-2 | 27 689 (57) | 27.8 (23.3-33.1) | 8 964 (6 796-11 490) | 92 (82-101) |
|  | 3+ | 10 403 (21) | 27.7 (23.1-32.9) | 9 080 (6 863-11 606) | 91 (81-101) |
|  | Missing | 13 (0) | 29.6 (27.4-35.4) | 9 048 (8 602-12 202) | 91 (87-101) |
| Had menopause | No | 13 166 (27) | 30.0 (25.2-35.6) | 9 304 (7 094-11 913) | 95 (85-105) |
|  | Yes | 28 028 (58) | 27.1 (22.7-32.1) | 9 022 (6 838-11 550) | 91 (81-101) |
|  | Missing/Not sure | 7 284 (15) | 26.8 (22.3-31.9) | 8 406 (6 290-10 923) | 89 (79-100) |
| Hormone-replacement therapy ever use | No | 31 109 (64) | 28.5 (24.0-34.0) | 9 215 (7 016-11 805) | 94 (83-103) |
|  | Yes | 17 283 (36) | 26.4 (22.1-31.3) | 8 612 (6 473-11 102) | 89 (79-99) |

| **Characteristic** |  | **No. (%)** | **Overall acceleration (mg)** | **Daily step count Median (IQR)** | **Peak 30 min cadence (steps/min) Median (IQR)** |
| --- | --- | --- | --- | --- | --- |
|  | Missing | 86 (0) | 26.2 (21.6-32.1) | 7 854 (6 521-10 788) | 87 (79-100) |
| Oral contraceptive pill ever use | No | 7 027 (14) | 26.9 (22.4-32.3) | 8 816 (6 685-11 295) | 91 (80-101) |
|  | Yes | 41 381 (85) | 27.9 (23.4-33.2) | 9 037 (6 835-11 591) | 92 (82-102) |
|  | Missing | 70 (0) | 29.2 (23.8-33.0) | 8 848 (6 806-11 994) | 90 (83-104) |

Activity metrics reported as median (interquartile range (IQR)). TDI = Townsend Deprivation Index, mg = milligravity, min=minute, cig = cigarettes, kg/m<sup>2</sup> = kilograms/meters<sup>2</sup>. <sup>a</sup>Among female participants, N=48 478.

### Total daily PA and cancer incidence

Higher total PA was associated with a lower risk of PA-related cancer (hazard ratio (HR) per 1 SD (8.3 milligravity (m*g*) units), 0.85 [95% confidence interval (CI) 0.81-0.89]) in the multivariable-adjusted models (Figure 1; eTable 5). Similar associations were found in models assessing quintiles of total PA (eTable 6).

To explore which cancers contributed to this result, we also examined associations for individual cancer sites (Figure 1; eTable 5). With higher daily PA, we observed a significantly lower risk for seven types; gastric cardia (HR_1SD_=0.39, [95% CI 0.22-0.70]), liver (HR_1SD_=0.65, [0.47-0.90]), bladder (HR_1SD_=0.69, [0.55-0.87]), lung (HR_1SD_=0.75, [0.65-0.86]), endometrial (HR_1SD_=0.78, [0.65-0.93]), colon (HR_1SD_=0.84, [0.75-0.95]), and breast cancers (HR_1SD_=0.91, [0.85-0.97]). There were also suggestive associations (HR < 0.9) for three cancers; oesophageal adenocarcinoma (HR_1SD_=0.89, [0.68-1.16]), kidney (HR_1SD_=0.86, [0.71-1.04]), and head and neck cancers (HR_1SD_=0.84, [0.66-1.07]). Similar associations were found in models assessing quintiles of total PA (eTable 6).

In secondary analysis of non-PA-related cancer sites, we observed no protective association with prostate cancer, an inverse association with non-Hodgkin lymphoma (HR_1SD_=0.85, [0.75-0.97]), and suggestive inverse associations for melanoma (HR_1SD_=0.89, [0.79-1.01]) and pancreatic cancers (HR_1SD_=0.86 [0.71-1.04]).

### Compositional data analysis

First, we estimated the risks associated with each behaviour individually by reallocating time to one behaviour from all others proportionally for an average individual (eFigure 2). Reallocating one hour/day to LIPA from all other behaviours was associated with a 5% lower risk of PA-related cancer (HR=0.95, [0.93-0.97]) in multivariable-adjusted models. Reallocating one hour/day to MVPA from other behaviours was associated with a 6% lower cancer risk (HR=0.93, [0.90-0.97]). Reallocating one hour/day to sedentary behaviour (SB) was associated with a significantly higher cancer risk (HR=1.03, [1.01-1.06]), but reallocating one hour/day to sleep was not (HR=1.01, [0.98-1.05]).

**Figure 2.**
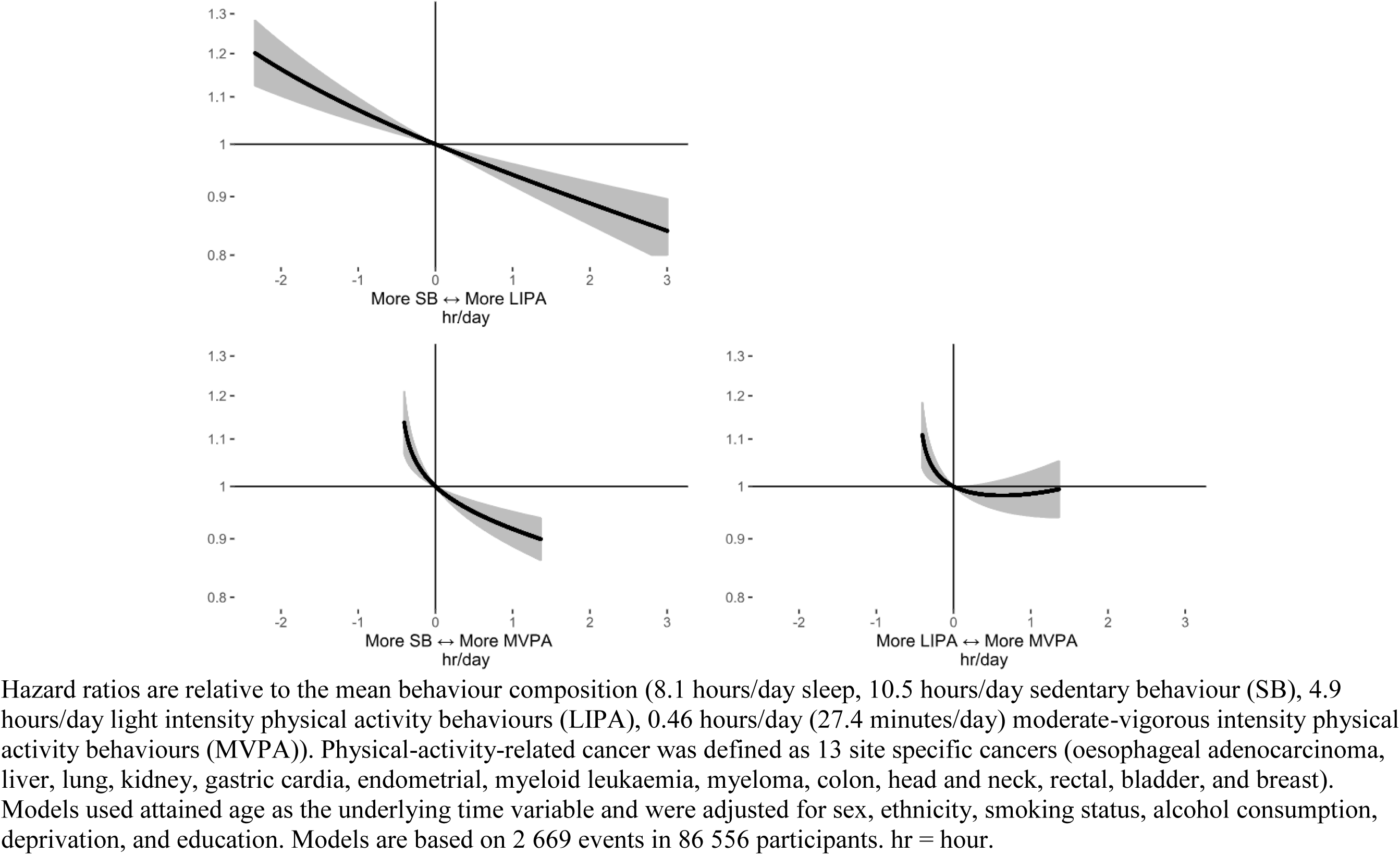
Hazard ratios for balance between movement behaviours and incident physical-activity-related cancer risk associated in 86 556 UK Biobank participants.

Next, we estimated the risks associated with specific pairwise reallocations of time between behaviours (Figure 2; eTable 7). For an average participant, reallocating one hour/day from SB to overall physical activity (LIPA + MVPA) was associated with a 7% lower risk (HR=0.93, [0.91-0.95]) in the multivariable-adjusted model. Reallocating one hour/day from SB to MVPA alone was associated with an 8% lower cancer risk (HR=0.92, [0.89-0.95]), and reallocating one hour/day from SB to LIPA was associated with a 6% lower risk (HR=0.94, [0.92-0.96]).

### Step counts and cancer incidence

Higher daily step counts were associated with a lower risk of PA-related cancers (*P* for trend≤0.001) after multivariable adjustments (eTable 8; Figure 3). Compared to individuals who took 5 000 daily steps (10^th^ percentile, reference), individuals who took 9 000 daily steps had an 18% lower risk (HR=0.82, [0.74-0.90]), while those who took 13 000 steps had a 23% lower risk (HR=0.77, [0.69-0.86]). Individuals taking fewer than 5 000 steps had a higher risk.

**Figure 3.**
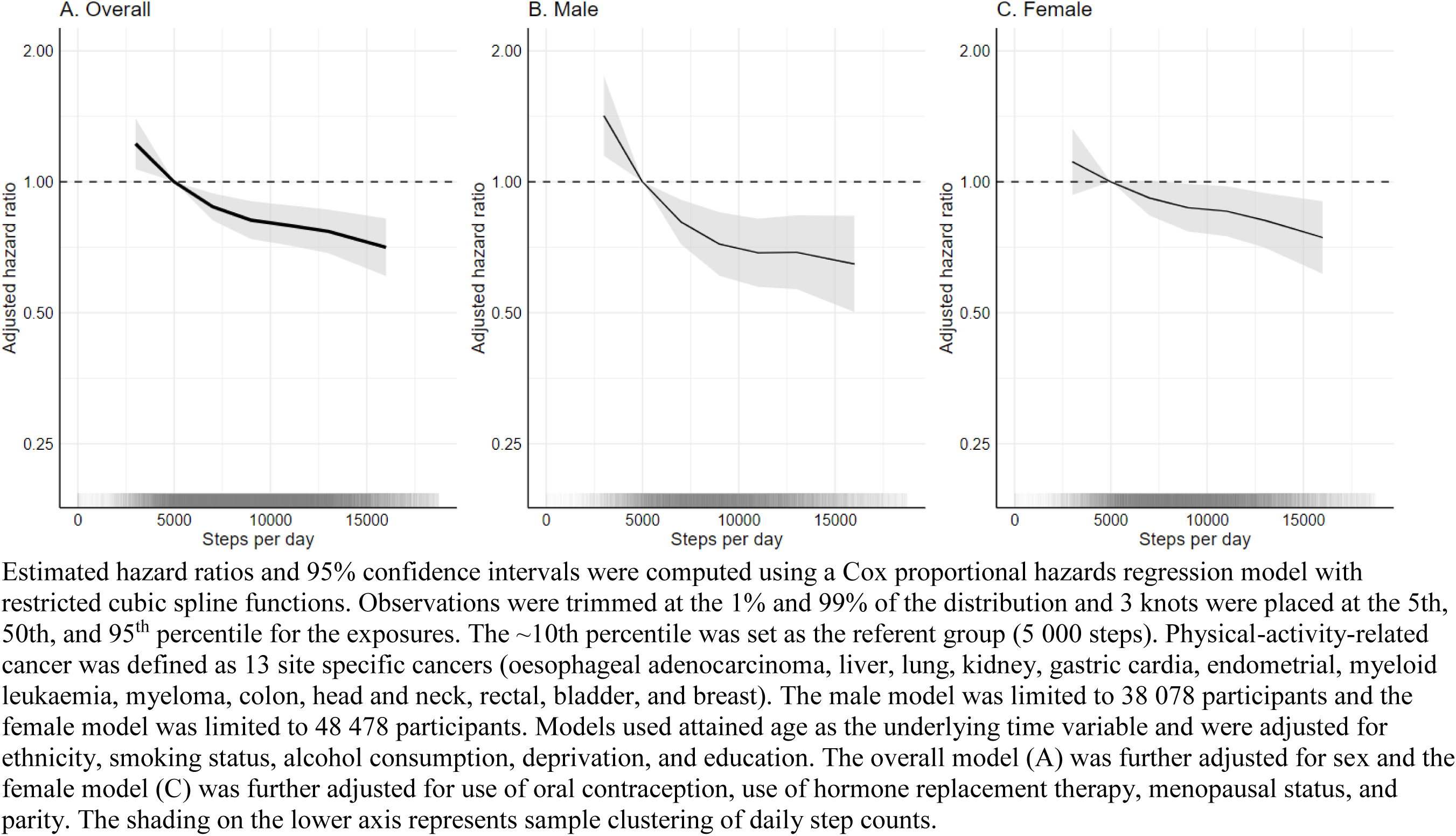
Dose-response associations between daily step count and physical-activity-related cancer risk in 86 556 UK Biobank participants.

Higher step intensity (peak 30-minute cadence) was associated with a lower risk of PA-related cancer prior to adjustment for total step counts (Table 2). Compared to individuals who took 70 steps/minute (10^th^ percentile, reference), individuals who took 120 steps/minute had a lower cancer risk before (HR=0.72, [0.59-0.89]), but not after adjusting for total daily steps (HR=0.88, [0.69-1.14], *P* for trend=0.05).

**Table 2.** Adjusted hazard ratios for peak 30-minute cadence and physical activity-related cancer risk in 86 556 UK Biobank participants.

|  | **Model 1** | **Model 2** |
| --- | --- | --- |
| **Peak 30-minute cadence (steps/minute)** | **HR (95% CI)** | **HR (95% CI)** |
| 60 | 1.14 (1.02 - 1.27) | 1.10 (0.98 - 1.23) |
| 70 (REF, ~10<sup>th</sup> percentile) | 1.00 (1.00 - 1.00) | 1.00 (1.00 - 1.00) |
| 80 | 0.88 (0.80 - 0.96) | 0.91 (0.83 - 1.00) |
| 90 | 0.82 (0.73 - 0.92) | 0.89 (0.78 - 1.01) |
| 100 | 0.77 (0.69 - 0.87) | 0.88 (0.76 - 1.02) |
| 110 | 0.74 (0.65 - 0.84) | 0.87 (0.73 - 1.04) |
| 120 (~95<sup>th</sup> percentile) | 0.72 (0.59 - 0.89) | 0.88 (0.69 - 1.14) |
| *P* value for linear trend | <0.001 | 0.05 |

Estimated hazard ratios (HRs) and 95% confidence intervals (CIs) were computed using a Cox proportional hazards regression model with restricted cubic spline functions. Observations were trimmed at the 1% and 99% of the distribution and 3 knots were placed at the 5th, 50th, and 95<sup>th</sup> percentile for the exposures. The ~10th percentile was set as the referent (REF) group (70 steps per minute).
Models used age as the underlying time variable and were adjusted for sex, ethnicity, smoking status, alcohol consumption, deprivation, and education. Model 2: Model 1 + median daily step count. Physical activity-related cancer was defined as 13 site specific cancers (oesophageal adenocarcinoma, liver, lung, kidney, gastric cardia, endometrial, myeloid leukaemia, myeloma, colon, head and neck, rectal, bladder, and breast).

### Sensitivity analyses

Associations for total PA and cancer risk were slightly attenuated after adjusting for body mass index (BMI) in models per 1-SD of total activity (eTable 5), but the direction between the quintiles was consistent (eTable 6). BMI adjustment did not substantially alter results from the compositional analyses (eFigure 3; eTable 7) or the spline models for step count (eTable 8), nor did adjusting for dietary factors (eTables 5-8, eFigure 4).

Sex-specific models of 1-SD differences in total PA showed that patterns of association for cancer risk were similar among males and females for most cancer sites (eTables 9-10), as were findings from the compositional data analysis (eFigure 5). Protective associations were observed for step count and PA-related cancer risk in sex-specific models (eTable 11; Figure 3). Analyses conducted among never smokers were similar to our primary analysis (eTable 12; eFigure 6). After removing the first two years of follow-up, higher activity quintiles demonstrated protective associations for cancer risk, but the impact was more attenuated in lower activity categories (eTable 13; eFigure 7).

## DISCUSSION

In this prospective analysis, higher total daily physical activity (PA) measured by accelerometers was associated with a composite of 13 cancers previously shown to be associated with PA in studies of self-reported leisure time activity.^17^ We also observed lower risks for 7 of these 13 cancers. Additionally, we found protective associations for minimizing sedentary time in favour of engaging in light intensity physical activity (LIPA) or moderate-vigorous physical activity (MVPA). Compared with individuals who took 5 000 steps per day, those who took 9 000 daily steps had an 18% lower risk of incident PA-related cancer. A weaker suggestive association with higher step intensity was observed, but results were not significant after adjusting for total step count.

Results from our compositional data analysis are novel and suggest that less sedentary time in favour of LIPA or MVPA was associated with a lower risk of certain cancers. These findings align with previous studies indicating that sedentary behaviour may be associated with chronic disease risk factors and cancer development and progression.^27–29^ Notably, our finding that any intensity of PA was beneficial contrasts with findings from cardiovascular disease research, where intensity plays a more pivotal role in determining health benefits.^7,27^ A study by Stamatakis found that short bursts of vigorous intensity PA were associated with a lower risk of PA-related cancer among non-exercising adults.^30^ Our findings suggests that efforts to decrease sedentary behaviour in favour of engaging in LIPA or MVPA activities, such as allocating more time to casual walking, household chores, home repairs, and gardening could also support cancer prevention efforts and might be feasible to incorporate into everyday routines. Further, our study was not limited to non-exercisers, which enhances the applicability of our findings.

Walking, a highly accessible and popular form of PA, is often considered an ideal exercise intervention due to its simplicity to track and minimal adverse effects.^31^ Our findings show that higher steps counts were associated with a lower risk for PA-related cancers. In terms of step intensity, we found a weaker but non-significant association after adjusting for total step count. These findings align with our compositional model results for total PA, which suggested that LIPA and MVPA were beneficial for cancer risk. Together, this suggests that walking at any pace may provide health benefits with respect to cancer incidence.

Our study adds new information to a limited literature base on the association between total daily PA and incident cancers.^11^ Previous research in the UK Biobank cohort reported protective associations for self-reported PA and cancer, and we extend these findings for many site-specific cancers.^28,32^ Our findings align with other studies using accelerometer data that found physical activity may play a role in the prevention of PA-related cancers among older women^33^ and for incident breast cancer.^15,16^ In addition to finding lower risk for 7 of the 13 PA-related cancers identified by Moore and colleagues,^17^ we also observed a protective association between total PA and non-Hodgkin lymphoma and suggestive associations for melanoma and pancreatic cancers. This suggests potential areas for future research, especially in samples with larger case counts.

Several hypothesized mechanisms linking higher PA with a lower cancer risk have been proposed. These mechanisms encompass hormonal changes, insulin levels, inflammation, immune function, and oxidative stress.^29^ Some hypotheses suggest that PA may reduce cancer risk by influencing body weight, but we observed no substantial changes in model estimates after adjusting for body mass index, although measurements here were obtained several years before the accelerometer measurements, potentially introducing imprecise estimates over time.

### Strengths and limitations

Our study has several strengths, including the use of accelerometer devices in a large and prospective cohort study, which reduces susceptibility to recall and reporting biases compared to most previous studies.^3,4^ Accelerometers captured a wide range of daily behaviours, including both sedentary and physically active activities, as well as step count. Further, we processed the accelerometer data using open-source methods.^34^ Our sample included a wide age-range (43-78 years at accelerometer wear) of participants, making the cohort mature for analysing more common adult-onset cancers.^19,35^ Our results add to the limited literature on how physical activity impacts cancer risk across different cancer sites. We also employed compositional data analyses to model associations for 24-hour behaviours, rather than just individual behaviour risks. In addition, we controlled for key cancer risk factors and conducted extensive sensitivity analysis to examine major threats to the validity of our findings.

This study also had several limitations. The UK Biobank cohort includes middle to early late-aged individuals, potentially leading to underrepresentation of cancers with onset at more advanced ages. Secondly, the study is observational, and we cannot exclude the possibility of unmeasured and residual confounding. Reverse causation is also a possibility, as certain cancers may have been undiagnosed at the time of accelerometer wear and could have reduced daily PA. Nonetheless after removing the first two years of follow-up, we observed only minor attenuations. Future UK Biobank studies with additional follow-up will be required to investigate associations for less common cancers. Furthermore, we had accelerometer data from one period during middle-age; thus, we are also unable to draw conclusions about PA earlier in the life in relation to cancer risk. Finally, the study population drawn from the UK Biobank may not be representative of wider populations.^36^ However, we believe that the etiological findings are still likely to have broader applicability.^37^

## Conclusion

Results from this prospective study suggest protective associations between engaging in higher levels of overall daily PA, reducing sedentary time in favour of engaging LIPA or MVPA, and increasing daily step counts and cancer risk. These findings underscore the potential health benefits of incorporating lower intensity activities into daily life during middle age, alongside promoting higher intensity activities in public health initiatives focused on cancer prevention.

## Data Availability

We do have permission to share the data. However, all bona fide researchers can apply to use the UK Biobank resource for health-related research that is in the public interest (https://www.ukbiobank.ac.uk/register-apply/).

## Author Contributions

Shreves had full access to all the data in the study and takes responsibility for the integrity of the data and the accuracy of the data analysis.

Concept and design: Shreves, Travis, Matthews, Doherty.

Acquisition, analysis, or interpretation of data: Shreves, Small, Walmsley, Saint-Maurice, Papier, Travis, Matthews, Doherty.

Drafting of the manuscript: Shreves, Travis, Matthews, Doherty.

Critical revision of the manuscript for important intellectual content: All authors.

Statistical analysis: Shreves.

Obtained funding: Shreves, Travis, Matthews, Doherty.

Administrative, technical, or material support: Shreves, Chan, Walmsley, Doherty.

Supervision: Doherty, Matthews, Travis.

## Conflict of Interest Disclosures

Shreves, Moore, Gaitskell, Travis, and Matthews declare no competing interests.

Doherty is supported by Novo Nordisk. Doherty accepted consulting fees from the University of Wisconsin (NIH R01 grant) and Harvard University (NIH R01 grant). Doherty received support for presentations at the International Conference on Soft Computing in Data Science 2021, Korean Society of Cardiology 2020, and Oxford International Study Abroad programme 2023. Doherty received support to attend the 2023 CTSU Joint Annual Collaborators’ Meeting, London, 2023 Big Model AI for Drug Design workshop, Abu Dhabi, 2023 Festival of Genomics, London, 2022 Industry Symposium on Digital Biomarkers, EMBL-EBI, Cambridge, 2020 ELIXIR Bioinformatics Industry Forum, London, 2020 3rd NYU Biomedical and Biosystems Conference, Abu Dhabi, 2020 Academy of Medical Sciences, UK-Japan Symposium on Data-Driven Health. Doherty is a member of the 2021 Scientific advisory board EU IMI IDEA-FAST. Doherty received a donation from SwissRe to purchase equipment for accelerometer data collection in the China Kadoorie Biobank.

Doherty, Walmsley, and Chan have released accelerometer analysis software under an academic use license that could result in commercial entities paying license fees for their use.

## Funding/Support

Shreves, Saint-Maurice, Moore, and Matthews are supported by the National Institutes of Health’s Intramural Research Program. Shreves is also supported by the National Institutes of Health’s Oxford Cambridge Scholars Program. Walmsley is supported by HDR UK, an initiative funded by UK Research and Innovation, Department of Health and Social Care (England) and the devolved administrations. Papier is supported by Cancer Research UK (grant number C16077/A29186). Chan is supported Novo Nordisk. Doherty and Small are supported by the Wellcome Trust (223100/Z/21/Z). Doherty is also supported by Novo Nordisk, Swiss Re, and the British Heart Foundation Centre of Research Excellence (grant number RE/18/3/34214). Travis is supported by Cancer Research UK (grant number C8221/A29017).

## Role of the Funder/Sponsor

The funders/sponsors had no role in the design of this study, the analysis and interpretation of the results, or drafting of the manuscript.

## eTABLES

**eTable 1:**
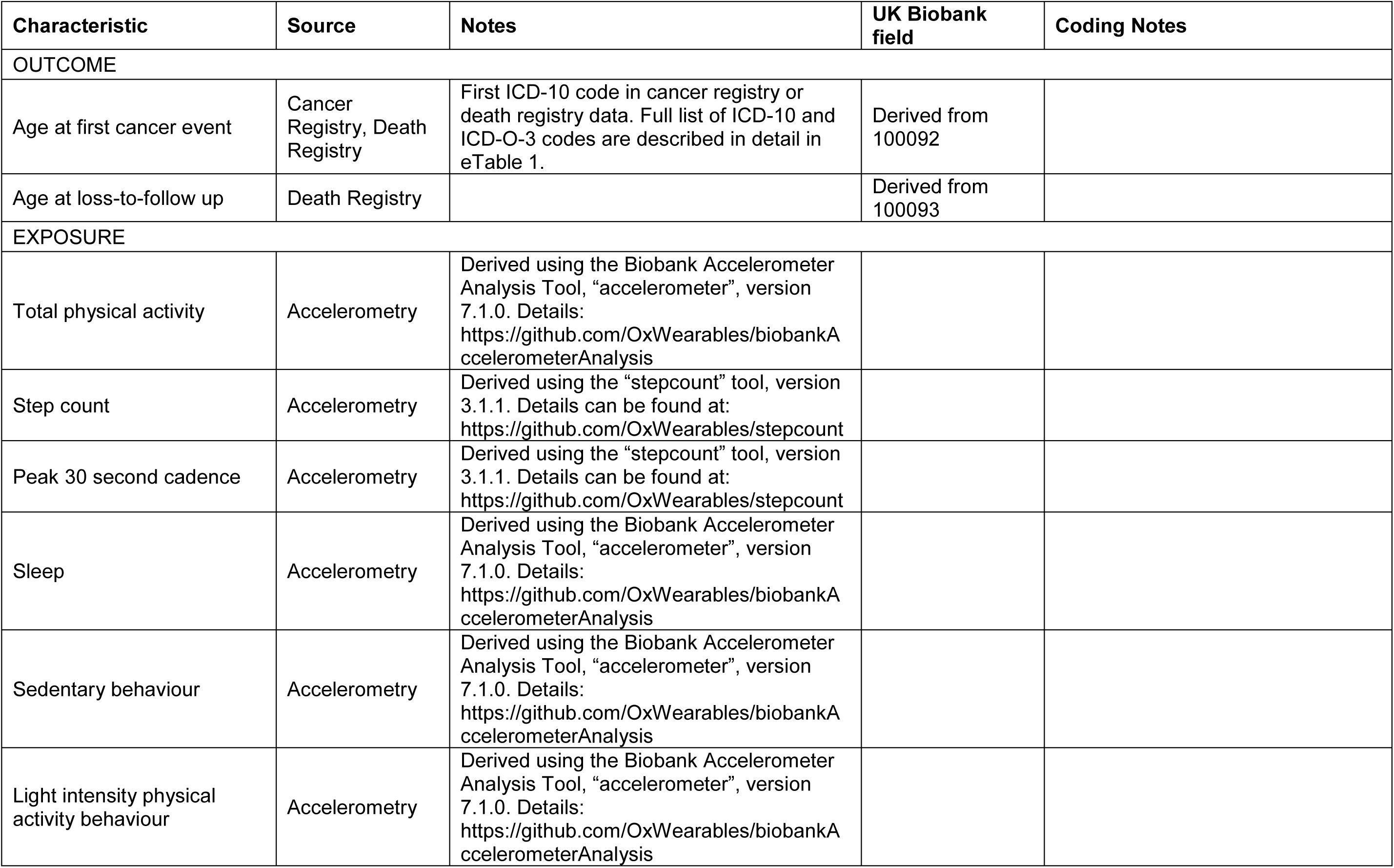
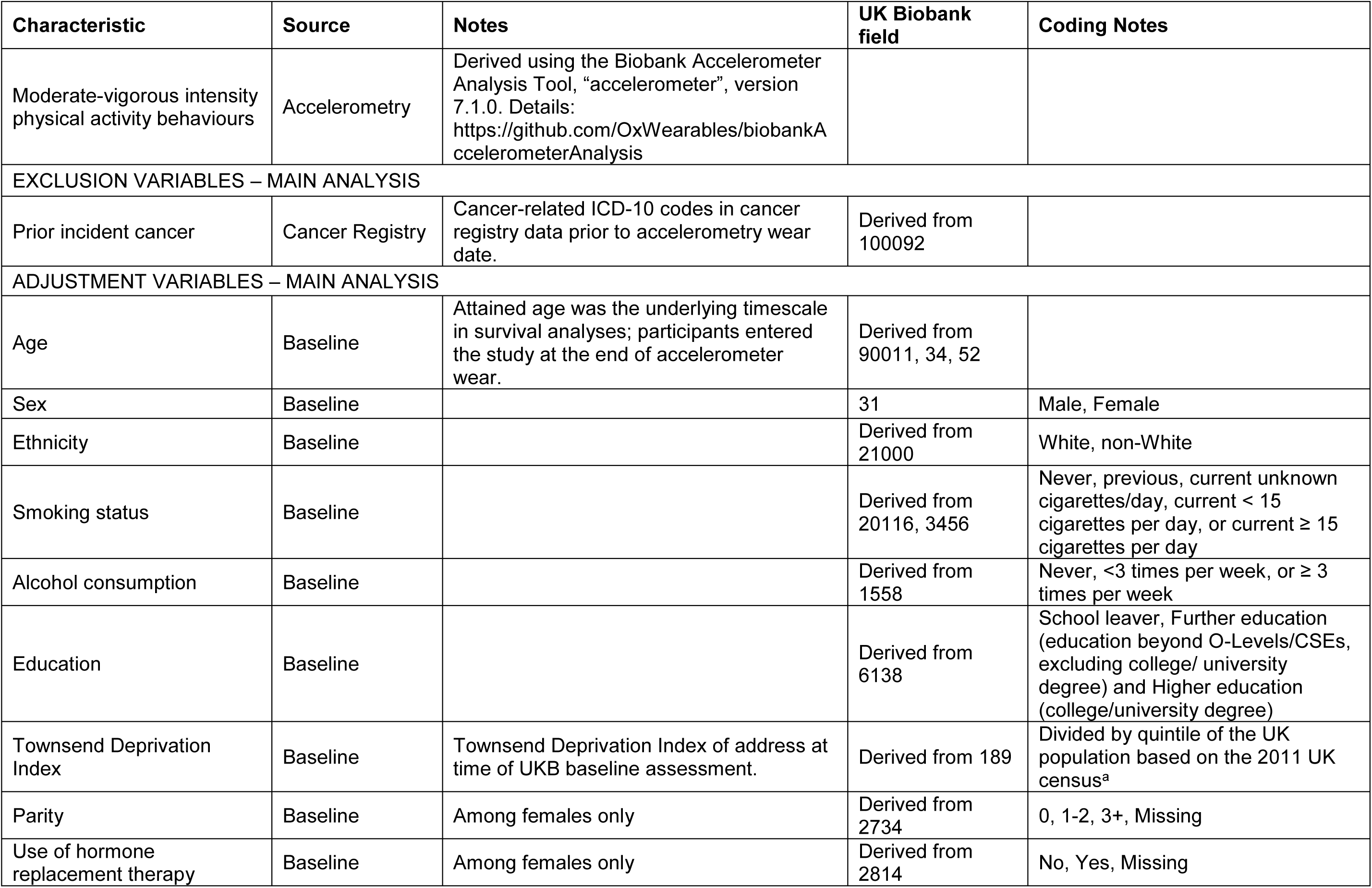
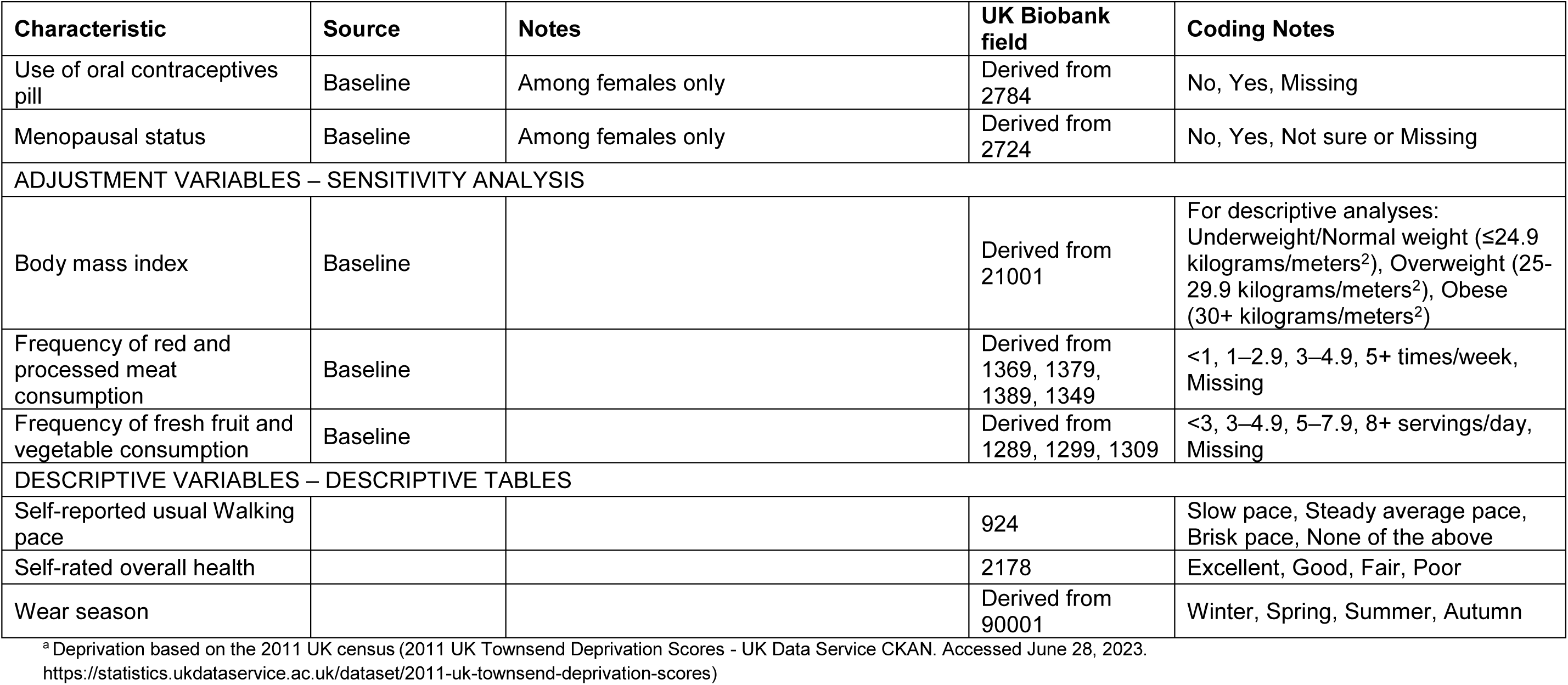
Definition of variables from the UK Biobank data.

| Characteristic | Source | Notes | UK Biobank field | Coding Notes |
| --- | --- | --- | --- | --- |
| OUTCOME |  |  |  |  |
| Age at first cancer event | Cancer Registry, Death Registry | First ICD-10 code in cancer registry or death registry data. Full list of ICD-10 and ICD-O-3 codes are described in detail in eTable 1. | Derived from 100092 |  |
| Age at loss-to-follow up | Death Registry |  | Derived from 100093 |  |
| EXPOSURE |  |  |  |  |
| Total physical activity | Accelerometry | Derived using the Biobank Accelerometer Analysis Tool, “accelerometer”, version 7.1.0. Details: https://github.com/OxWearables/biobankAccelerometerAnalysis |  |  |
| Step count | Accelerometry | Derived using the “stepcount” tool, version 3.1.1. Details can be found at: https://github.com/OxWearables/stepcount |  |  |
| Peak 30 second cadence | Accelerometry | Derived using the “stepcount” tool, version 3.1.1. Details can be found at: https://github.com/OxWearables/stepcount |  |  |
| Sleep | Accelerometry | Derived using the Biobank Accelerometer Analysis Tool, “accelerometer”, version 7.1.0. Details: https://github.com/OxWearables/biobankAccelerometerAnalysis |  |  |
| Sedentary behaviour | Accelerometry | Derived using the Biobank Accelerometer Analysis Tool, “accelerometer”, version 7.1.0. Details: https://github.com/OxWearables/biobankAccelerometerAnalysis |  |  |
| Light intensity physical activity behaviour | Accelerometry | Derived using the Biobank Accelerometer Analysis Tool, “accelerometer”, version 7.1.0. Details: https://github.com/OxWearables/biobankAccelerometerAnalysis |  |  |
| Moderate-vigorous intensity physical activity behaviours | Accelerometry | Derived using the Biobank Accelerometer Analysis Tool, “accelerometer”, version 7.1.0. Details: https://github.com/OxWearables/biobankAccelerometerAnalysis |  |  |
| EXCLUSION VARIABLES – MAIN ANALYSIS |  |  |  |  |
| Prior incident cancer | Cancer Registry | Cancer-related ICD-10 codes in cancer registry data prior to accelerometry wear date. | Derived from 100092 |  |
| ADJUSTMENT VARIABLES – MAIN ANALYSIS |  |  |  |  |
| Age | Baseline | Attained age was the underlying timescale in survival analyses; participants entered the study at the end of accelerometer wear. | Derived from 90011, 34, 52 |  |
| Sex | Baseline |  | 31 | Male, Female |
| Ethnicity | Baseline |  | Derived from 21000 | White, non-White |
| Smoking status | Baseline |  | Derived from 20116, 3456 | Never, previous, current unknown cigarettes/day, current < 15 cigarettes per day, or current ≥ 15 cigarettes per day |
| Alcohol consumption | Baseline |  | Derived from 1558 | Never, <3 times per week, or ≥ 3 times per week |
| Education | Baseline |  | Derived from 6138 | School leaver, Further education (education beyond O-Levels/CSEs, excluding college/ university degree) and Higher education (college/university degree) |
| Townsend Deprivation Index | Baseline | Townsend Deprivation Index of address at time of UKB baseline assessment. | Derived from 189 | Divided by quintile of the UK population based on the 2011 UK census<sup>a</sup> |
| Parity | Baseline | Among females only | Derived from 2734 | 0, 1-2, 3+, Missing |
| Use of hormone replacement therapy | Baseline | Among females only | Derived from 2814 | No, Yes, Missing |
| Use of oral contraceptives pill | Baseline | Among females only | Derived from 2784 | No, Yes, Missing |
| Menopausal status | Baseline | Among females only | Derived from 2724 | No, Yes, Not sure or Missing |
| ADJUSTMENT VARIABLES – SENSITIVITY ANALYSIS |  |  |  |  |
| Body mass index | Baseline | | Derived from 21001 | For descriptive analyses: Underweight/Normal weight ($\leq 24.9$ kilograms/meters<sup>2</sup>), Overweight (25–29.9 kilograms/meters<sup>2</sup>), Obese (30+ kilograms/meters<sup>2</sup>) |
| Frequency of red and processed meat consumption | Baseline |  | Derived from 1369, 1379, 1389, 1349 | <1, 1–2.9, 3–4.9, 5+ times/week, Missing |
| Frequency of fresh fruit and vegetable consumption | Baseline |  | Derived from 1289, 1299, 1309 | <3, 3–4.9, 5–7.9, 8+ servings/day, Missing |
| DESCRIPTIVE VARIABLES – DESCRIPTIVE TABLES |  |  |  |  |
| Self-reported usual Walking pace |  |  | 924 | Slow pace, Steady average pace, Brisk pace, None of the above |
| Self-rated overall health |  |  | 2178 | Excellent, Good, Fair, Poor |
| Wear season |  |  | Derived from 90001 | Winter, Spring, Summer, Autumn |

<sup>a</sup> Deprivation based on the 2011 UK census (2011 UK Townsend Deprivation Scores - UK Data Service CKAN. Accessed June 28, 2023. https://statistics.ukdataservice.ac.uk/dataset/2011-uk-townsend-deprivation-scores)

**eTable 2.**
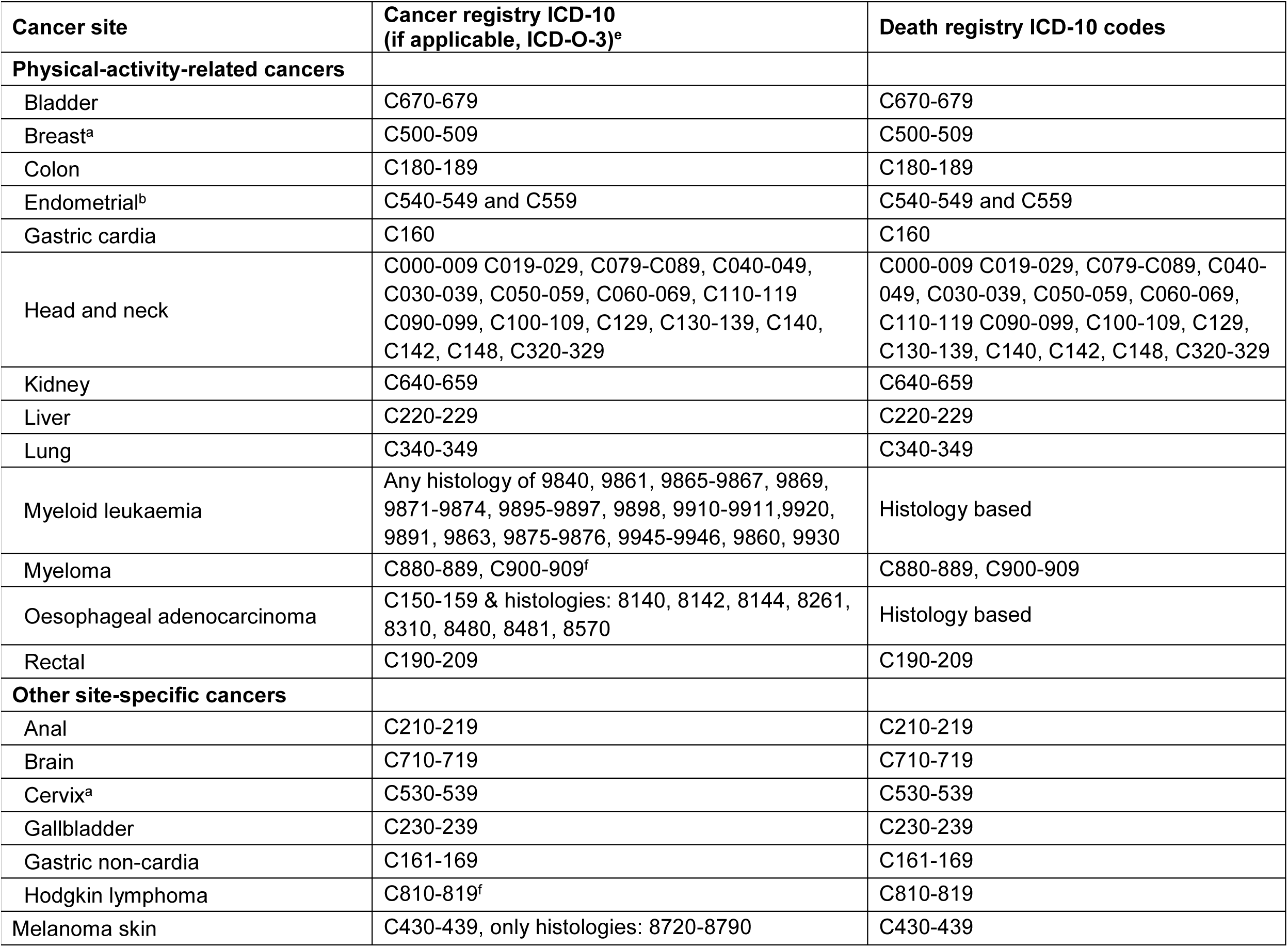
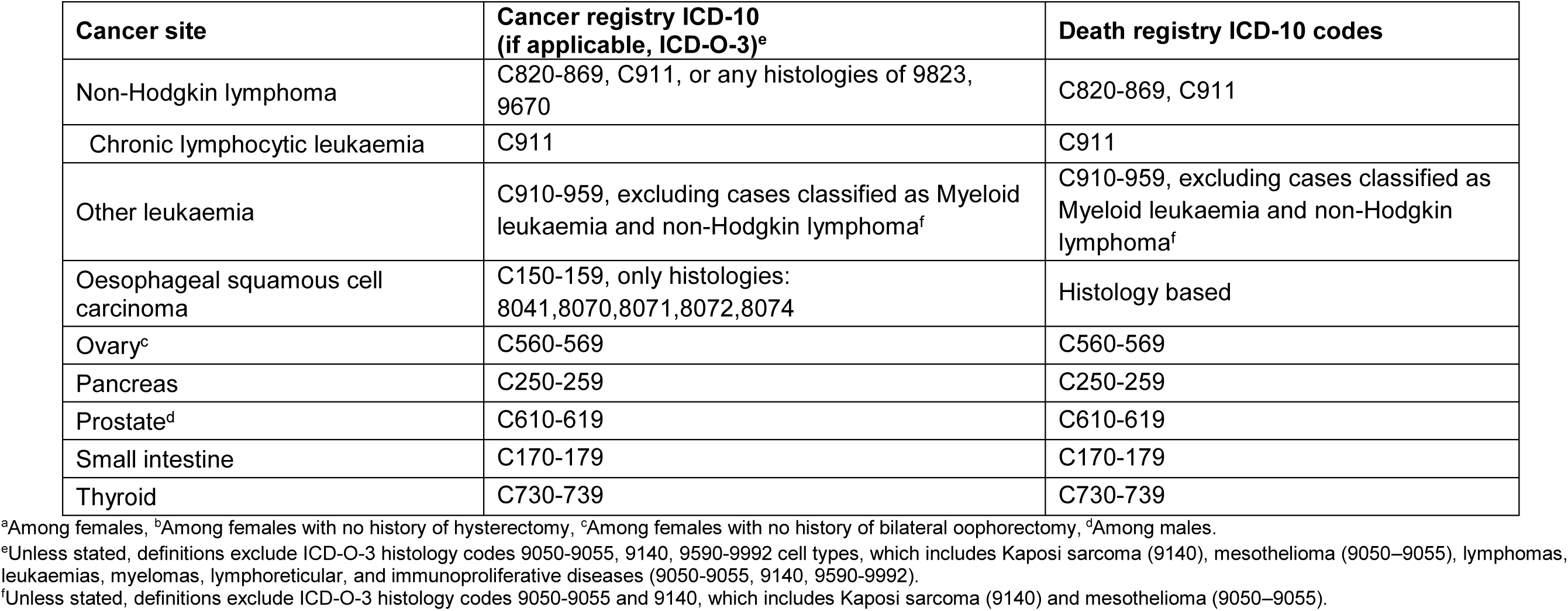
Coding and number of site-specific cancer incident cases in UK Biobank participants.

**eTable 3.** Tabulation of number of incident primary events per cancer site in 86 556 UK Biobank participants.

|  | Overall | Male | Female |
| --- | --- | --- | --- |
| **Cancer type** | **N=86 556** | **N=38 078** | **N=48 478** |
| **Physical-activity-related cancers** | 2 669 | 898 | 1 771 |
| Bladder | 109 | 86 | 23 |
| Breast<sup>a</sup> | 995 | NA | 995 |
| Colon | 381 | 207 | 174 |
| Endometrial<sup>b</sup> | 153 | NA | 153 |
| Gastric cardia | 22 | 18 | <5<sup>a</sup> |
| Head and neck | 88 | 68 | 20 |
| Kidney | 141 | 83 | 58 |
| Liver | 60 | 34 | 26 |
| Lung | 295 | 148 | 147 |
| Myeloid leukaemia | 43 | 18 | 25 |
| Myeloma | 107 | 61 | 46 |
| Oesophageal adenocarcinoma | 68 | 56 | 12 |
| Rectal | 209 | 121 | 88 |
| **Other site-specific cancers** |  |  |  |
| Anal | 26 | 6 | 20 |
| Brain | 79 | 42 | 37 |
| Cervix<sup>a</sup> | 12 | NA | 12 |
| Gallbladder | 10 | 6 | <5<sup>a</sup> |
| Gastric non-cardia | 40 | 25 | 15 |
| Hodgkin lymphoma | 12 | 7 | 5 |
| Melanoma skin | 328 | 181 | 147 |
| Non-Hodgkin lymphoma | 316 | 176 | 140 |
| Chronic lymphocytic leukaemia | 76 | 49 | 27 |
| Oesophageal squamous cell carcinoma | 29 | 12 | 17 |
| Other leukaemia | 27 | 18 | 9 |
| Ovary<sup>c</sup> | 66 | NA | 66 |
| Pancreas | 145 | 80 | 65 |
| Prostate<sup>d</sup> | 1380 | 1380 | NA |
| Small intestine | 30 | 14 | 16 |
| Thyroid | 43 | 15 | 28 |

Physical-activity-related cancer was defined as 13 site specific cancers (oesophageal adenocarcinoma, liver, lung, kidney, gastric cardia, endometrial, myeloid leukaemia, myeloma, colon, head and neck, rectal, bladder, and breast). <sup>a</sup>Among females, <sup>b</sup>Among females with no history of hysterectomy, <sup>c</sup>Among males, <sup>d</sup>Among females with no history of bilateral oophorectomy. <sup>e</sup>UK Biobank policy states that tables should have a minimum number of 5 reported participants within a cell.

**eTable 4.** Strengthening the Reporting of Observational Studies in Epidemiology (STROBE) statement.

|  | Item No. | Recommendation | Page No. |
| --- | --- | --- | --- |
| Title and abstract | 1 | (a) Indicate the study's design with a commonly used term in the title or the abstract | 1, 3 |
|  |  | (b) Provide in the abstract an informative and balanced summary of what was done and what was found | 3 |
| Introduction |  |  |  |
| Background/rationale | 2 | Explain the scientific background and rationale for the investigation being reported | 5-6 |
| Objectives | 3 | State specific objectives, including any prespecified hypotheses | 5-6 |
| Methods |  |  |  |
| Study design | 4 | Present key elements of study design early in the paper | 5-9 |
| Setting | 5 | Describe the setting, locations, and relevant dates, including periods of recruitment, exposure, follow-up, and data collection | 6-7 |
| Participants | 6 | (a) *Cohort study*—Give the eligibility criteria, and the sources and methods of selection of participants. Describe methods of follow-up<br>*Case-control study*—Give the eligibility criteria, and the sources and methods of case ascertainment and control selection. Give the rationale for the choice of cases and controls<br>*Cross-sectional study*—Give the eligibility criteria, and the sources and methods of selection of participants | 6-8 |
|  |  | (b) *Cohort study*—For matched studies, give matching criteria and number of exposed and unexposed<br>*Case-control study*—For matched studies, give matching criteria and the number of controls per case | NA |
| Variables | 7 | Clearly define all outcomes, exposures, predictors, potential confounders, and effect modifiers. Give diagnostic criteria, if applicable | 6-9, eTable 1, eTable 2 |
| Data sources/measurement | 8* | For each variable of interest, give sources of data and details of methods of assessment (measurement). Describe comparability of assessment methods if there is more than one group | 6-9, eTable 1 |
| Bias | 9 | Describe any efforts to address potential sources of bias | 9 |
| Study size | 10 | Explain how the study size was arrived at | 6-8, eFigure 1 |
| Quantitative variables | 11 | Explain how quantitative variables were handled in the analyses. If applicable, describe which groupings were chosen and why | 6-9, eTable 1 |
| Statistical methods | 12 | (a) Describe all statistical methods, including those used to control for confounding | 8-10 |
|  |  | (b) Describe any methods used to examine subgroups and interactions | 9 |
|  |  | (c) Explain how missing data were addressed | 6-9, eTable1, eFigure 1 |
|  |  | (d) *Cohort study*—If applicable, explain how loss to follow-up was addressed<br>*Case-control study*—If applicable, explain how matching of cases and controls was addressed<br>*Cross-sectional study*—If applicable, describe analytical methods taking account of sampling strategy | 8, eFigure 1 |
|  |  | (e) Describe any sensitivity analyses | 9 |
| Participants | 13* | (a) Report numbers of individuals at each stage of study—eg numbers potentially eligible, examined for eligibility, confirmed eligible, included in the study, completing follow-up, and analysed | eFigure 1 |
|  |  | (b) Give reasons for non-participation at each stage | eFigure 1 |
|  |  | (c) Consider use of a flow diagram | eFigure 1 |
| Descriptive data | 14* | (a) Give characteristics of study participants (eg demographic, clinical, social) and information on exposures and potential confounders | Table 1 |
|  |  | (b) Indicate number of participants with missing data for each variable of interest | Table 1 |
|  |  | (c) *Cohort study*—Summarise follow-up time (eg, average and total amount) | 10 |
| Outcome data | 15* | *Cohort study*—Report numbers of outcome events or summary measures over time | 10, eTable 3 |
|  |  | *Case-control study*—Report numbers in each exposure category, or summary measures of exposure | NA |
|  |  | *Cross-sectional study*—Report numbers of outcome events or summary measures | NA |

|  | **Item No.** | **Recommendation** | **Page No.** |
| --- | --- | --- | --- |
| Main results | 16 | (a) Give unadjusted estimates and, if applicable, confounder-adjusted estimates and their precision (eg, 95% confidence interval). Make clear which confounders were adjusted for and why they were included | Table 2, eTables 5-13. |
|  |  | (b) Report category boundaries when continuous variables were categorized | eTable 6, eTable 12, eTable 13 |
|  |  | (c) If relevant, consider translating estimates of relative risk into absolute risk for a meaningful time period | NA |
| Other analyses | 17 | Report other analyses done—eg analyses of subgroups and interactions, and sensitivity analyses | 12-13 |
| **Discussion** |  |  |  |
| Key results | 18 | Summarise key results with reference to study objectives | 10-12 |
| Limitations | 19 | Discuss limitations of the study, taking into account sources of potential bias or imprecision. Discuss both direction and magnitude of any potential bias | 15-16 |
| Interpretation | 20 | Give a cautious overall interpretation of results considering objectives, limitations, multiplicity of analyses, results from similar studies, and other relevant evidence | 16 |
| Generalisability | 21 | Discuss the generalisability (external validity) of the study results | 15-16 |
| **Other information** |  |  |  |
| Funding | 22 | Give the source of funding and the role of the funders for the present study and, if applicable, for the original study on which the present article is based | 19-20 |

**eTable 5.**
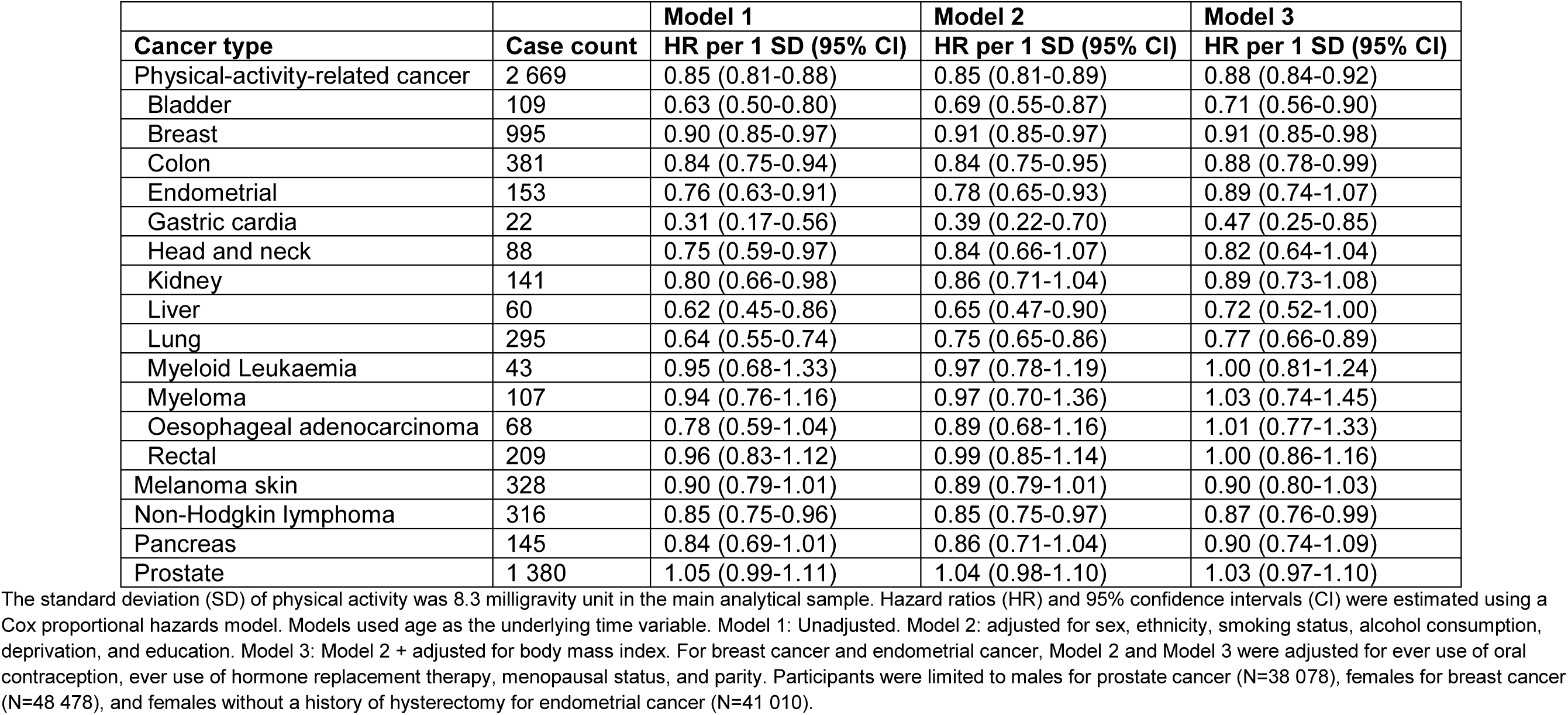
Sequential model adjustments for total physical activity (milligravity units) and risk of incident cancer in 86 556 UK Biobank participants.

|  |  | **Model 1** | **Model 2** | **Model 3** |
| --- | --- | --- | --- | --- |
| **Cancer type** | **Case count** | **HR per 1 SD (95% CI)** | **HR per 1 SD (95% CI)** | **HR per 1 SD (95% CI)** |
| Physical-activity-related cancer | 2 669 | 0.85 (0.81-0.88) | 0.85 (0.81-0.89) | 0.88 (0.84-0.92) |
| Bladder | 109 | 0.63 (0.50-0.80) | 0.69 (0.55-0.87) | 0.71 (0.56-0.90) |
| Breast | 995 | 0.90 (0.85-0.97) | 0.91 (0.85-0.97) | 0.91 (0.85-0.98) |
| Colon | 381 | 0.84 (0.75-0.94) | 0.84 (0.75-0.95) | 0.88 (0.78-0.99) |
| Endometrial | 153 | 0.76 (0.63-0.91) | 0.78 (0.65-0.93) | 0.89 (0.74-1.07) |
| Gastric cardia | 22 | 0.31 (0.17-0.56) | 0.39 (0.22-0.70) | 0.47 (0.25-0.85) |
| Head and neck | 88 | 0.75 (0.59-0.97) | 0.84 (0.66-1.07) | 0.82 (0.64-1.04) |
| Kidney | 141 | 0.80 (0.66-0.98) | 0.86 (0.71-1.04) | 0.89 (0.73-1.08) |
| Liver | 60 | 0.62 (0.45-0.86) | 0.65 (0.47-0.90) | 0.72 (0.52-1.00) |
| Lung | 295 | 0.64 (0.55-0.74) | 0.75 (0.65-0.86) | 0.77 (0.66-0.89) |
| Myeloid Leukaemia | 43 | 0.95 (0.68-1.33) | 0.97 (0.78-1.19) | 1.00 (0.81-1.24) |
| Myeloma | 107 | 0.94 (0.76-1.16) | 0.97 (0.70-1.36) | 1.03 (0.74-1.45) |
| Oesophageal adenocarcinoma | 68 | 0.78 (0.59-1.04) | 0.89 (0.68-1.16) | 1.01 (0.77-1.33) |
| Rectal | 209 | 0.96 (0.83-1.12) | 0.99 (0.85-1.14) | 1.00 (0.86-1.16) |
| Melanoma skin | 328 | 0.90 (0.79-1.01) | 0.89 (0.79-1.01) | 0.90 (0.80-1.03) |
| Non-Hodgkin lymphoma | 316 | 0.85 (0.75-0.96) | 0.85 (0.75-0.97) | 0.87 (0.76-0.99) |
| Pancreas | 145 | 0.84 (0.69-1.01) | 0.86 (0.71-1.04) | 0.90 (0.74-1.09) |
| Prostate | 1 380 | 1.05 (0.99-1.11) | 1.04 (0.98-1.10) | 1.03 (0.97-1.10) |

The standard deviation (SD) of physical activity was 8.3 milligravity unit in the main analytical sample. Hazard ratios (HR) and 95% confidence intervals (CI) were estimated using a Cox proportional hazards model. Models used age as the underlying time variable. Model 1: Unadjusted. Model 2: adjusted for sex, ethnicity, smoking status, alcohol consumption, deprivation, and education. Model 3: Model 2 + adjusted for body mass index. For breast cancer and endometrial cancer, Model 2 and Model 3 were adjusted for ever use of oral contraception, ever use of hormone replacement therapy, menopausal status, and parity. Participants were limited to males for prostate cancer (N=38 078), females for breast cancer (N=48 478), and females without a history of hysterectomy for endometrial cancer (N=41 010).

**eTable 6.** Sequential model adjustments for quintile of total physical activity (milligravity units), quintile of median daily step count, and risk of incident physical-activity-related cancer in 86 556 UK Biobank participants.

|  |  |  | Model 1 | Model 2 | Model 3 | Model 4 |
| --- | --- | --- | --- | --- | --- | --- |
| Quintile of total daily physical activity | Total | Case count | HR (95% CI) | HR (95% CI) | HR (95% CI) | HR (95% CI) |
| Q1: <21.6 | 17 311 | 718 | REF | REF | REF | REF |
| Q2: 21.6-25.4 | 17 311 | 569 | 0.84 (0.75-0.93) | 0.84 (0.75-0.93) | 0.86 (0.77-0.96) | 0.84 (0.75-0.94) |
| Q3: 25.5-29.1 | 17 311 | 522 | 0.80 (0.71-0.89) | 0.79 (0.71-0.89) | 0.83 (0.74-0.93) | 0.80 (0.71-0.90) |
| Q4: 29.2-34.2 | 17 311 | 460 | 0.74 (0.65-0.83) | 0.74 (0.65-0.83) | 0.78 (0.69-0.88) | 0.75 (0.66-0.84) |
| Q5: 34.3+ | 17 312 | 400 | 0.69 (0.61-0.78) | 0.70 (0.61-0.79) | 0.76 (0.66-0.86) | 0.71 (0.62-0.80) |
| *P* value for linear trend |  |  | <0.001 | <0.001 | <0.001 | <0.001 |
| Quintile of daily step count | Total | Case count | HR (95% CI) | HR (95% CI) | HR (95% CI) | HR (95% CI) |
| Q1: < 6 319 | 17 308 | 687 | REF | REF | REF | REF |
| Q2: 6 319-8 187 | 17 302 | 542 | 0.80 (0.71-0.90) | 0.82 (0.74-0.92) | 0.85 (0.76-0.95) | 0.85 (0.74-0.98) |
| Q3: 8 188-10 009 | 17 315 | 513 | 0.76 (0.68-0.85) | 0.79 (0.71-0.89) | 0.83 (0.74-0.93) | 0.83 (0.72-0.95) |
| Q4: 10 010-12 415 | 17 318 | 494 | 0.74 (0.66-0.83) | 0.78 (0.69-0.88) | 0.83 (0.73-0.93) | 0.79 (0.69-0.91) |
| Q5: 12 416+ | 17 313 | 433 | 0.66 (0.59-0.75) | 0.71 (0.63-0.80) | 0.76 (0.67-0.86) | 0.76 (0.66-0.88) |
| *P* value for linear trend |  |  | <0.001 | <0.001 | <0.001 | <0.001 |

Hazard ratios (HR) and 95% confidence intervals (CI) were estimated using a Cox proportional hazards model. Physical-activity-related cancer was defined as 13 site specific cancers (oesophageal adenocarcinoma, liver, lung, kidney, gastric cardia, endometrial, myeloid leukaemia, myeloma, colon, head and neck, rectal, bladder, and breast). Models used age as the underlying time variable. Model 1: Unadjusted. Model 2: adjusted for sex, ethnicity, smoking status, alcohol consumption, deprivation, and education. Model 3: Model 2 + adjusted for body mass index. Model 4: Model 2 + adjusted for fruit and vegetable intake, and red and processed meat intake. REF = reference.

**eTable 7.** Changes in behaviour time and physical-activity-related cancer risk in 86 556 UK Biobank participants.

|  | **Model 1** | **Model 2** | **Model 3** | **Model 4** |
| --- | --- | --- | --- | --- |
| **Change in behaviour time** | **HR (95% CI)** | **HR (95% CI)** | **HR (95% CI)** | **HR (95% CI)** |
| 1 hour/day to LIPA from SB | 0.97 (0.95-1.00) | 0.94 (0.92-0.96) | 0.95 (0.93-0.97) | 0.94 (0.92-0.96) |
| 1 hour/day to MVPA from SB | 0.87 (0.84-0.90) | 0.92 (0.89-0.95) | 0.94 (0.91-0.98) | 0.92 (0.89-0.95) |
| 1 hr/day to Any PA from SB | 0.94 (0.93-0.96) | 0.93 (0.91-0.95) | 0.94 (0.92-0.96) | 0.93 (0.91-0.95) |
| 1 hr/day to MVPA from LIPA | 0.90 (0.86-0.93) | 0.99 (0.94-1.03) | 1.00 (0.96-1.05) | 0.99 (0.94-1.03) |

Hazard ratios (HR) and 95% confidence intervals (CI) for incident physical-activity-related cancer associated with reallocating time to from one behaviour to another behaviour. Models are based on 2 669 events in 86 556 participants. HRs are relative to the mean behaviour composition (8.1 hours/day sleep, 10.5 hours/day sedentary behaviour (SB), 4.9 hours/day light intensity physical activity behaviours (LIPA), 0.46 hours/day (27.4 minutes/day) moderate-vigorous intensity physical (MVPA) activity behaviours). The "Any PA" category represents time in either LIPA or MVPA and was generated using a combined category for LIPA and MVPA activity time. Physical-activity-related cancer was defined as 13 site specific cancers (oesophageal adenocarcinoma, liver, lung, kidney, gastric cardia, endometrial, myeloid leukaemia, myeloma, colon, head and neck, rectal, bladder, and breast). Models used age as the underlying time variable. Model 1: Unadjusted. Model 2: adjusted for sex, ethnicity, smoking status, alcohol consumption, deprivation, and education. Model 3: Model 2 + adjusted for body mass index. Model 4: Model 2 + adjusted for fruit and vegetable intake, and red and processed meat intake. Models are based on 2 669 events in 86 556 participants.

**eTable 8.**
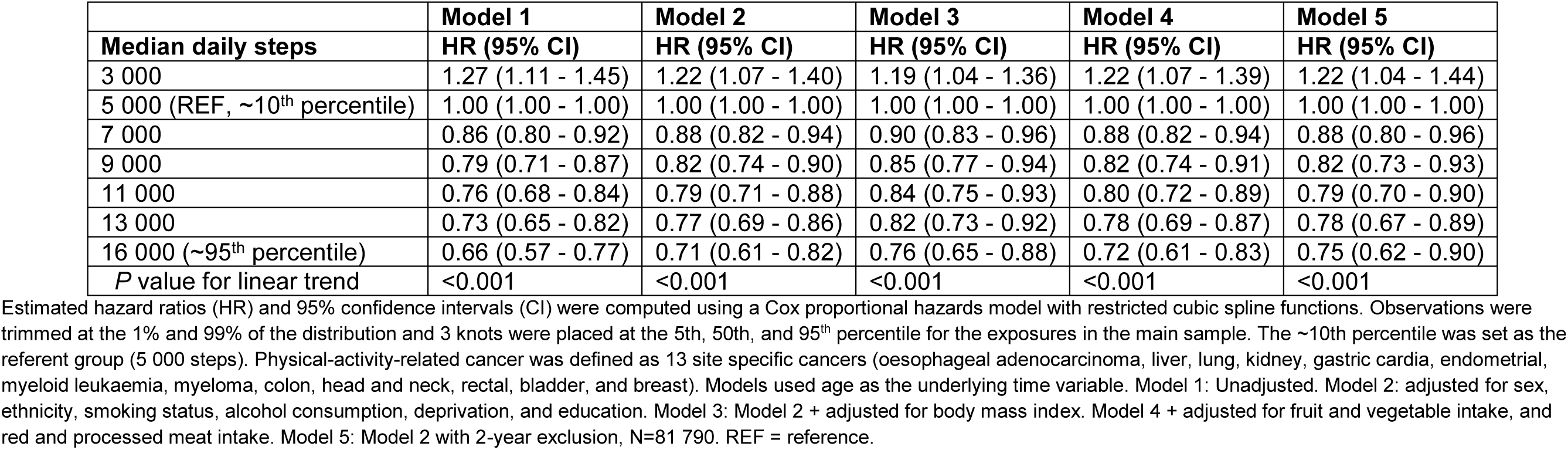
Sequential model adjustments for median daily step count and physical-activity-related cancer risk in 86 556 UK Biobank participants.

**eTable 9.** Sequential model adjustments for total physical activity (milligravity units) and risk of incident cancer in the UK Biobank among 38 078 male UK Biobank participants.

|  |  | **Model 1** | **Model 2** | **Model 3** |
| --- | --- | --- | --- | --- |
| **Cancer type** | **Case count** | **HR per 1 SD (95% CI)** | **HR per 1 SD (95% CI)** | **HR per 1 SD (95% CI)** |
| Physical-activity-related cancer | 898 | 0.77 (0.71-0.84) | 0.80 (0.74-0.87) | 0.84 (0.78-0.91) |
| Bladder | 86 | 0.70 (0.54-0.92) | 0.72 (0.56-0.95) | 0.76 (0.58-1.00) |
| Breast | NA | NA | NA | NA |
| Colon | 207 | 0.77 (0.65-0.91) | 0.76 (0.64-0.90) | 0.80 (0.67-0.95) |
| Endometrial | NA | NA | NA | NA |
| Gastric cardia | 18 | 0.34 (0.17-0.67) | 0.41 (0.21-0.79) | 0.49 (0.25-0.98) |
| Head and neck | 68 | 0.86 (0.65-1.13) | 0.91 (0.69-1.18) | 0.88 (0.67-1.16) |
| Kidney | 83 | 0.85 (0.66-1.09) | 0.89 (0.69-1.13) | 0.90 (0.70-1.16) |
| Liver | 34 | 0.76 (0.50-1.15) | 0.77 (0.51-1.17) | 0.84 (0.55-1.29) |
| Lung | 148 | 0.54 (0.43-0.68) | 0.64 (0.52-0.80) | 0.68 (0.54-0.84) |
| Myeloid Leukaemia | 18 | 1.04 (0.63-1.72) | 1.11 (0.67-1.82) | 1.26 (0.76-2.10) |
| Myeloma | 61 | 0.92 (0.69-1.22) | 0.93 (0.70-1.24) | 0.94 (0.70-1.27) |
| Oesophageal adenocarcinoma | 56 | 0.83 (0.61-1.13) | 0.89 (0.66-1.20) | 1.01 (0.74-1.38) |
| Rectal | 121 | 0.95 (0.78-1.16) | 0.96 (0.79-1.17) | 0.98 (0.80-1.20) |
| Melanoma skin | 181 | 0.90 (0.77-1.07) | 0.88 (0.75-1.05) | 0.90 (0.76-1.08) |
| Non-Hodgkin lymphoma | 176 | 0.91 (0.77-1.08) | 0.90 (0.75-1.07) | 0.91 (0.76-1.09) |
| Pancreas | 80 | 0.84 (0.64-1.09) | 0.86 (0.66-1.11) | 0.93 (0.71-1.21) |
| Prostate | 1 380 | 1.05 (0.99-1.11) | 1.04 (0.98-1.10) | 1.03 (0.97-1.10) |

The standard deviation (SD) of physical activity was 8.3 milligravity unit in the main analytical sample. Hazard ratios (HR) and 95% confidence intervals (CI) were estimated using a Cox proportional hazards model. Models used age as the underlying time variable. Model 1: Unadjusted. Model 2: adjusted for ethnicity, smoking status, alcohol consumption, deprivation, and education. Model 3: Model 2 + adjusted for body mass index.

**eTable 10.** Sequential model adjustments for total physical activity (milligravity units) and risk of incident cancer in the UK Biobank among 48 478 female UK Biobank participants.

|  |  | **Model 1** | **Model 2** | **Model 3** |
| --- | --- | --- | --- | --- |
| **Cancer type** | **Case count** | **HR per 1 SD (95% CI)** | **HR per 1 SD (95% CI)** | **HR per 1 SD (95% CI)** |
| Physical-activity-related cancer | 1 771 | 0.87 (0.83-0.92) | 0.88 (0.84-0.93) | 0.90 (0.86-0.95) |
| Bladder | 23 | 0.56 (0.33-0.94) | 0.54 (0.31-0.91) | 0.52 (0.30-0.90) |
| Breast | 995 | 0.90 (0.85-0.97) | 0.91 (0.85-0.97) | 0.91 (0.85-0.98) |
| Colon | 174 | 0.95 (0.80-1.12) | 0.94 (0.80-1.11) | 0.96 (0.81-1.14) |
| Endometrial | 153 | 0.76 (0.63-0.91) | 0.78 (0.65-0.93) | 0.89 (0.74-1.07) |
| Gastric cardia | <5<sup>a</sup> | NA | NA | NA |
| Head and neck | 20 | 0.55 (0.32-0.96) | 0.61 (0.36-1.05) | 0.59 (0.34-1.04) |
| Kidney | 58 | 0.78 (0.58-1.06) | 0.82 (0.61-1.11) | 0.88 (0.64-1.19) |
| Liver | 26 | 0.48 (0.29-0.80) | 0.50 (0.30-0.84) | 0.55 (0.32-0.93) |
| Lung | 147 | 0.75 (0.62-0.91) | 0.86 (0.71-1.04) | 0.87 (0.72-1.06) |
| Myeloid Leukaemia | 25 | 0.89 (0.57-1.38) | 0.89 (0.57-1.39) | 0.90 (0.57-1.42) |
| Myeloma | 46 | 0.99 (0.72-1.36) | 1.01 (0.74-1.39) | 1.08 (0.79-1.49) |
| Oesophageal adenocarcinoma | 12 | 0.80 (0.41-1.55) | 0.89 (0.46-1.72) | 1.02 (0.51-2.01) |
| Rectal | 88 | 1.02 (0.82-1.27) | 1.02 (0.82-1.28) | 1.03 (0.82-1.29) |
| Melanoma skin | 147 | 0.92 (0.77-1.09) | 0.90 (0.75-1.08) | 0.90 (0.75-1.09) |
| Non-Hodgkin lymphoma | 140 | 0.80 (0.66-0.97) | 0.81 (0.67-0.98) | 0.82 (0.67-1.00) |
| Pancreas | 65 | 0.86 (0.65-1.14) | 0.88 (0.66-1.17) | 0.87 (0.65-1.17) |
| Prostate | NA | NA | NA | NA |

The standard deviation (SD) of physical activity was 8.3 milligravity unit in the main analytical sample. Hazard ratios (HR) and 95% confidence intervals (CI) were estimated using a Cox proportional hazards model. Models used age as the underlying time variable. Model 1: Unadjusted. Model 2: Adjusted for ethnicity, smoking status, alcohol consumption, deprivation, education, ever use of oral contraception, ever use of hormone replacement therapy, menopausal status, and parity. Model 3: Model 2 + adjusted for body mass index. Participants were limited to females without a history of hysterectomy for endometrial cancer, N=41 010. <sup>a</sup>UK Biobank policy states that tables should have a minimum number of 5 reported participants within a cell.

**eTable 11.**
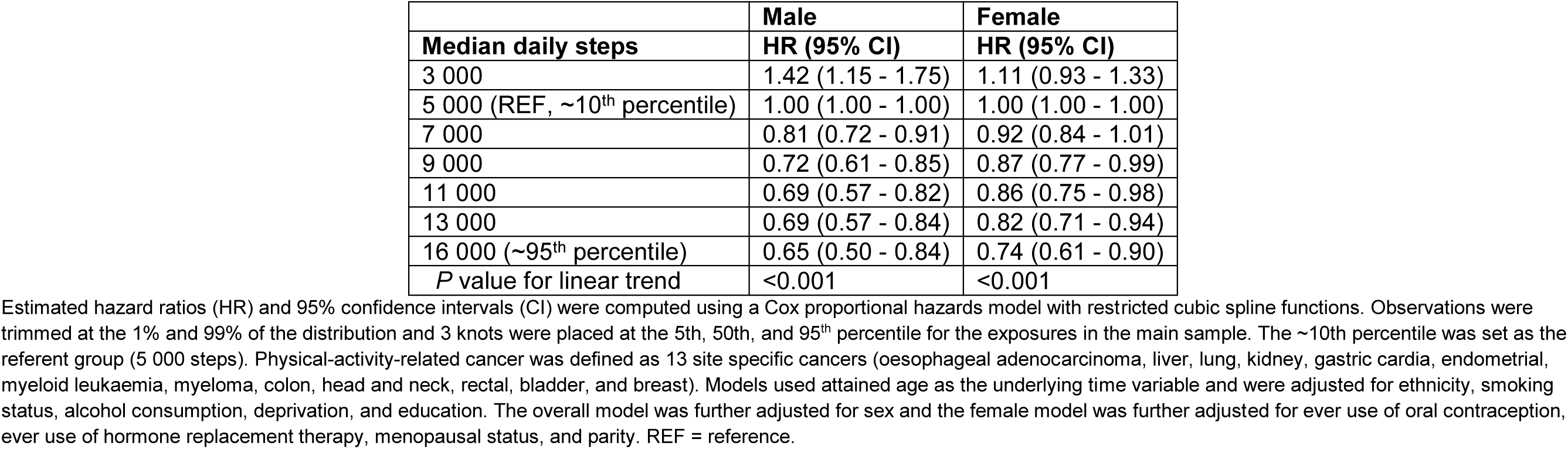
Sex-specific adjusted hazard ratios for median daily step count and physical-activity-related cancer risk in 38 078 male UK Biobank participants and 48 478 female UK Biobank participants.

**eTable 12.** Models for quintile of total physical activity (milligravity units), quintile of median daily step count, and risk of incident physical-activity-related cancer in UK Biobank participants who were never smokers (N=49 736).

| **Quintile of physical activity** | **Total** | **Case count** | **HR (95% CI)** |
| --- | --- | --- | --- |
| Q1: <22.0 | 9 947 | 335 | REF |
| Q2: 22.0-25.7 | 9 946 | 269 | 0.83 (0.70-0.97) |
| Q3: 25.8-29.5 | 9 948 | 265 | 0.83 (0.71-0.98) |
| Q4: 29.6-34.5 | 9 947 | 244 | 0.80 (0.67-0.94) |
| Q5: 34.6+ | 9 948 | 195 | 0.68 (0.57-0.82) |
| *P* value for linear trend |  |  | <0.001 |
| **Quintile of daily step count** | **Total** | **Case count** | **HR (95% CI)** |
| Q1: <6 480 | 9 941 | 308 | REF |
| Q2: 6 480-8 318 | 9 953 | 281 | 0.93 (0.79-1.09) |
| Q3: 8 319-10 109 | 9 943 | 253 | 0.84 (0.71-0.99) |
| Q4: 10 110-12 469 | 9 951 | 255 | 0.85 (0.72-1.01) |
| Q5: 12, 470+ | 9 948 | 211 | 0.73 (0.61-0.87) |
| *P* value for linear trend |  |  | 0.001 |

Physical-activity-related cancer was defined as 13 site specific cancers (oesophageal adenocarcinoma, liver, lung, kidney, gastric cardia, endometrial, myeloid leukaemia, myeloma, colon, head and neck, rectal, bladder, and breast). Hazard ratios (HR) and 95% confidence intervals (CI) were estimated using a Cox proportional hazards model. Models used age as the underlying time variable. Model was adjusted for sex, ethnicity, alcohol consumption, deprivation, and education. REF = reference.

**eTable 13.**
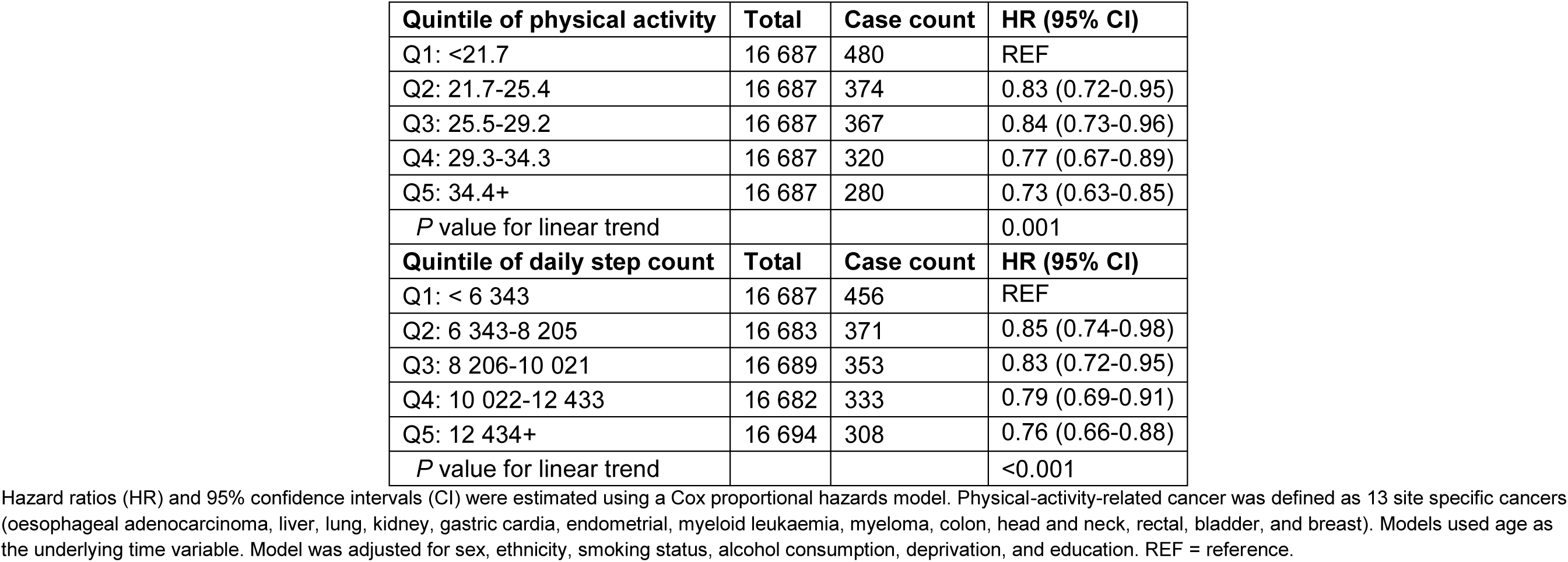
Models for quintile of total physical activity (milligravity units), quintile of median daily step count, and risk of incident physical-activity-related cancer in 86 556 UK Biobank participants before and after removing the first two years of follow-up (N=83 435).

| **Quintile of physical activity** | **Total** | **Case count** | **HR (95% CI)** |
| --- | --- | --- | --- |
| Q1: <21.7 | 16 687 | 480 | REF |
| Q2: 21.7-25.4 | 16 687 | 374 | 0.83 (0.72-0.95) |
| Q3: 25.5-29.2 | 16 687 | 367 | 0.84 (0.73-0.96) |
| Q4: 29.3-34.3 | 16 687 | 320 | 0.77 (0.67-0.89) |
| Q5: 34.4+ | 16 687 | 280 | 0.73 (0.63-0.85) |
| *P* value for linear trend |  |  | 0.001 |
| **Quintile of daily step count** | **Total** | **Case count** | **HR (95% CI)** |
| Q1: < 6 343 | 16 687 | 456 | REF |
| Q2: 6 343-8 205 | 16 683 | 371 | 0.85 (0.74-0.98) |
| Q3: 8 206-10 021 | 16 689 | 353 | 0.83 (0.72-0.95) |
| Q4: 10 022-12 433 | 16 682 | 333 | 0.79 (0.69-0.91) |
| Q5: 12 434+ | 16 694 | 308 | 0.76 (0.66-0.88) |
| *P* value for linear trend |  |  | <0.001 |

Hazard ratios (HR) and 95% confidence intervals (CI) were estimated using a Cox proportional hazards model. Physical-activity-related cancer was defined as 13 site specific cancers (oesophageal adenocarcinoma, liver, lung, kidney, gastric cardia, endometrial, myeloid leukaemia, myeloma, colon, head and neck, rectal, bladder, and breast). Models used age as the underlying time variable. Model was adjusted for sex, ethnicity, smoking status, alcohol consumption, deprivation, and education. REF = reference.

## eFIGURES

**eFigure 1.**
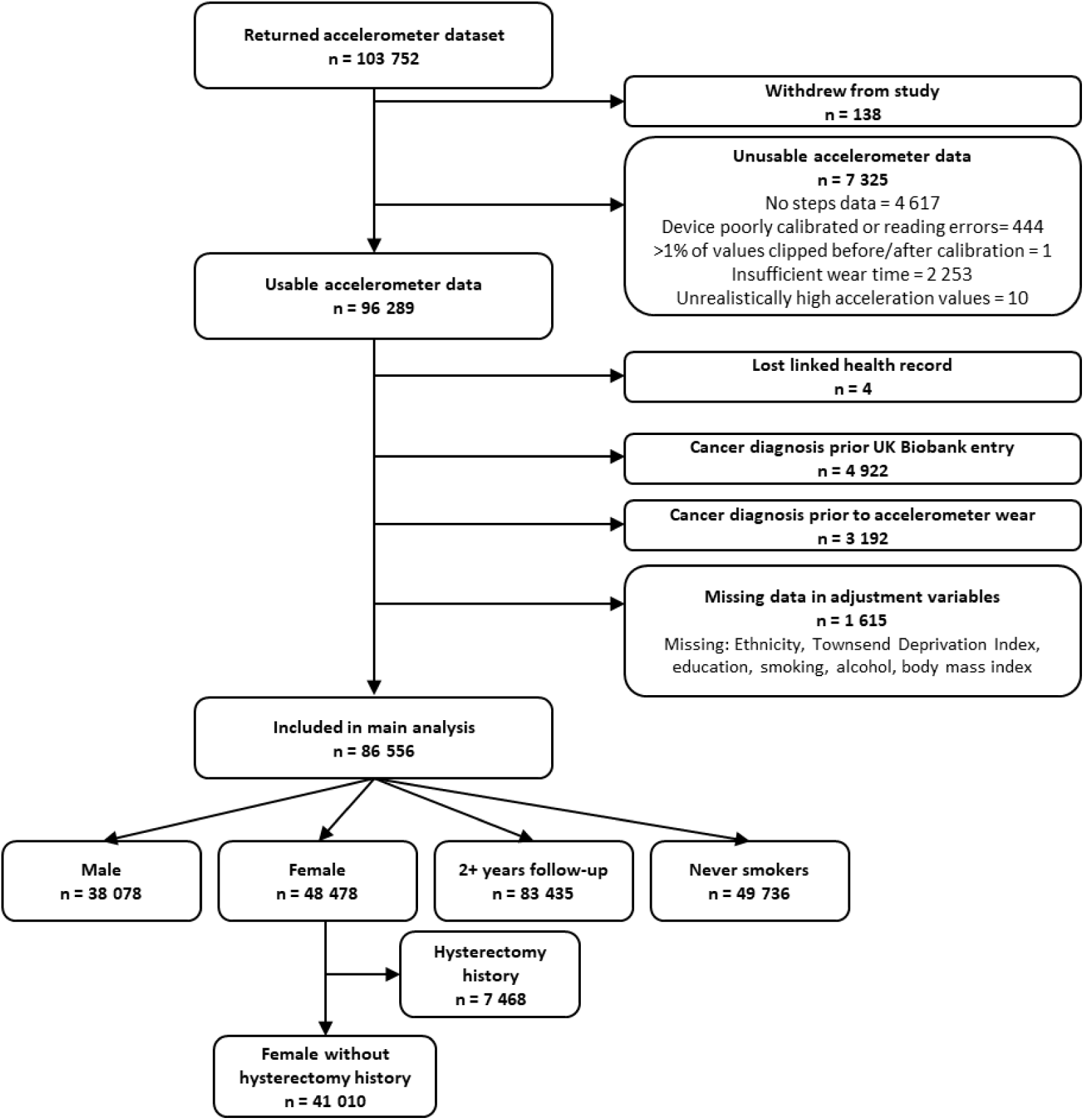
Participant flow diagram for the analysis of daily physical activity and step count measured by accelerometers in UK Biobank participants.

**eFigure 2.**
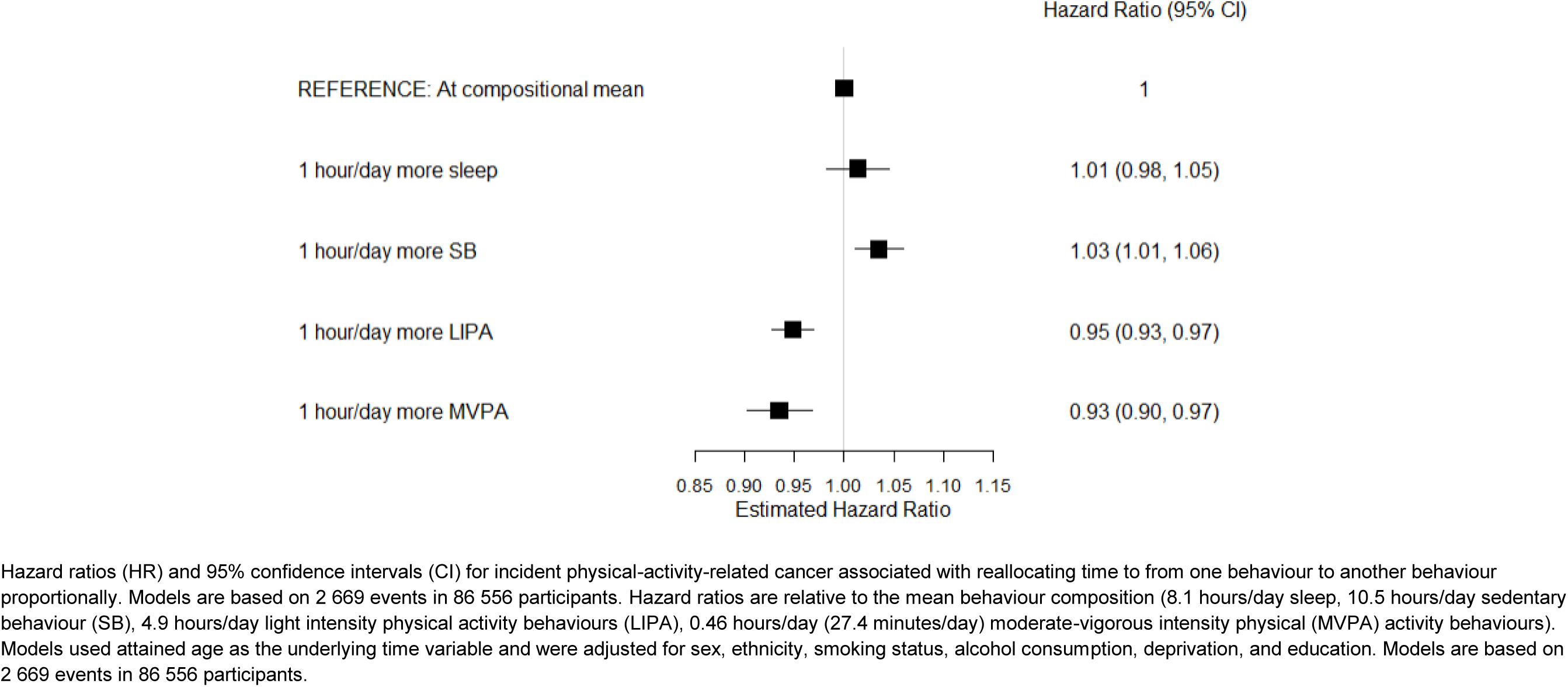
Hazard Ratios reallocating time to a given movement behaviour from all other behaviours proportionally and incident physical-activity-related cancer in 86 556 UK Biobank participants.

**eFigure 3.**
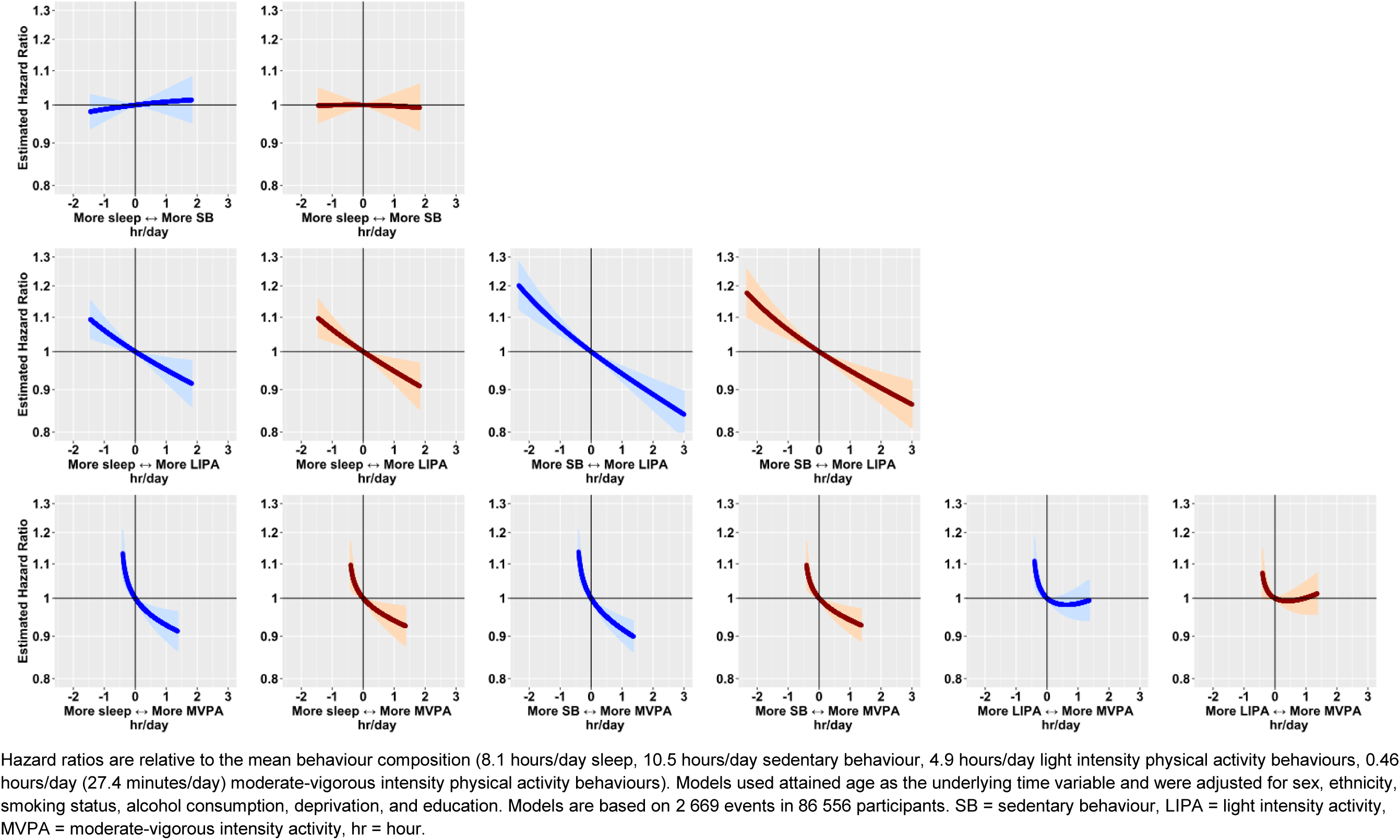
Hazard ratios for all behaviour pairs and incident physical-activity-related cancer risk for estimated using a multivariable-adjusted Cox regression model in 86 556 UK Biobank participants before (blue) and after adjusting for body mass index as a sensitivity analysis.

**eFigure 4.**
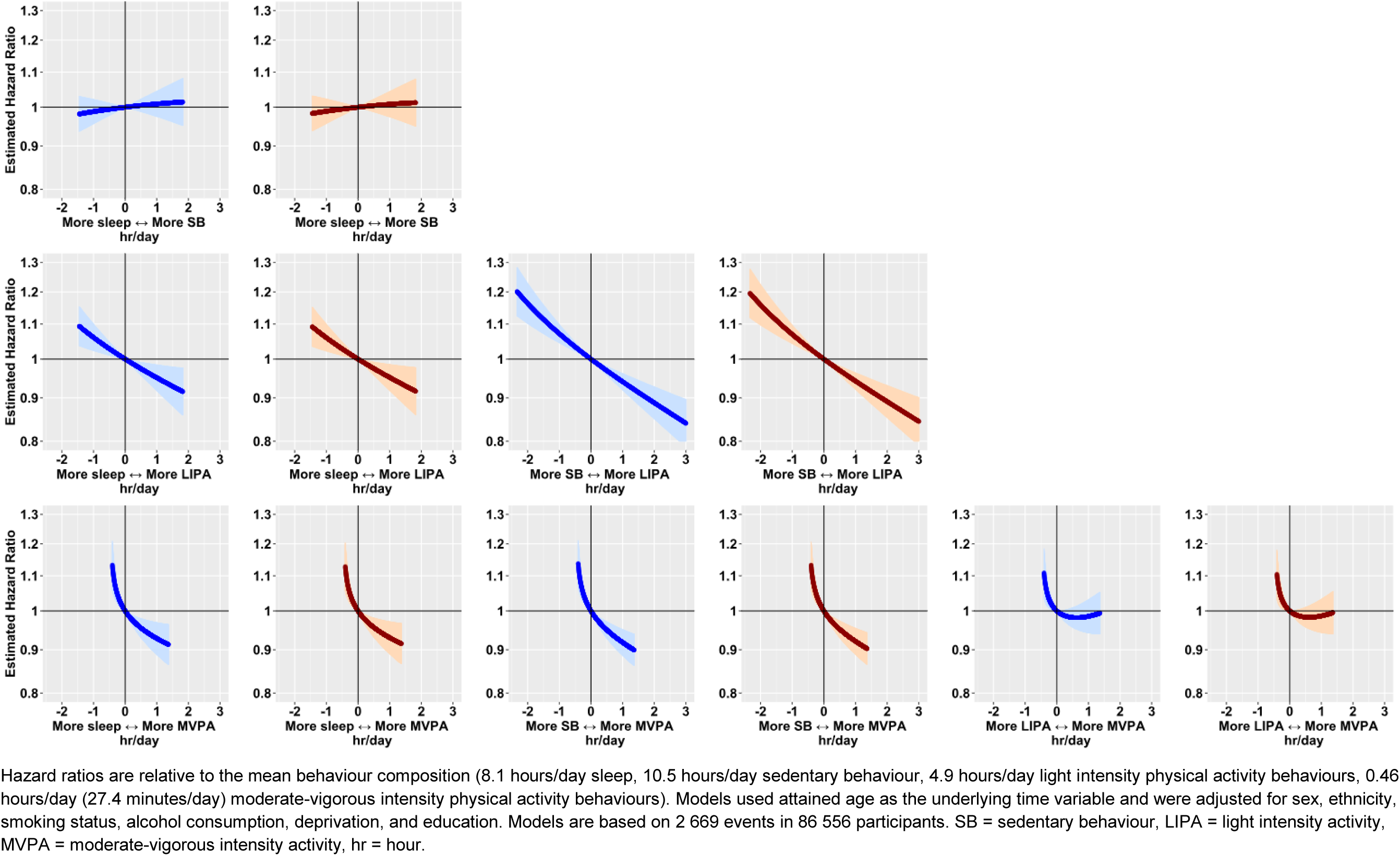
Hazard ratios for all behaviour pairs and incident physical-activity-related cancer risk for estimated using a multivariable-adjusted Cox regression model in 86 556 UK Biobank participants before (blue) and after adjusting for dietary factors as a sensitivity analysis.

**eFigure 5.**
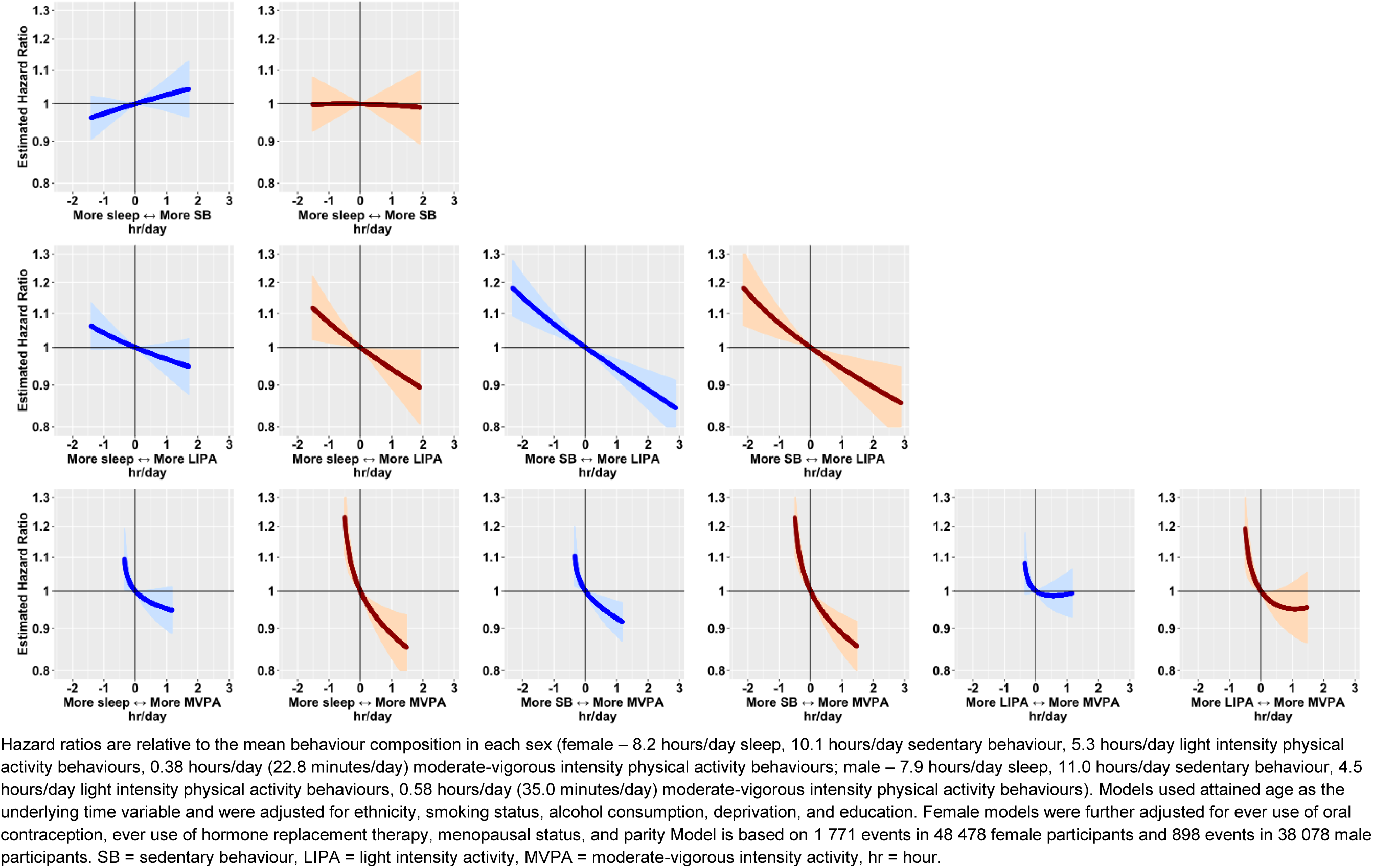
Hazard ratios for all behaviour pairs and incident physical-activity-related cancer risk for estimated using a multivariable-adjusted Cox regression model among 48 478 female participants (blue) and 38 078 male participants (red).

**eFigure 6.**
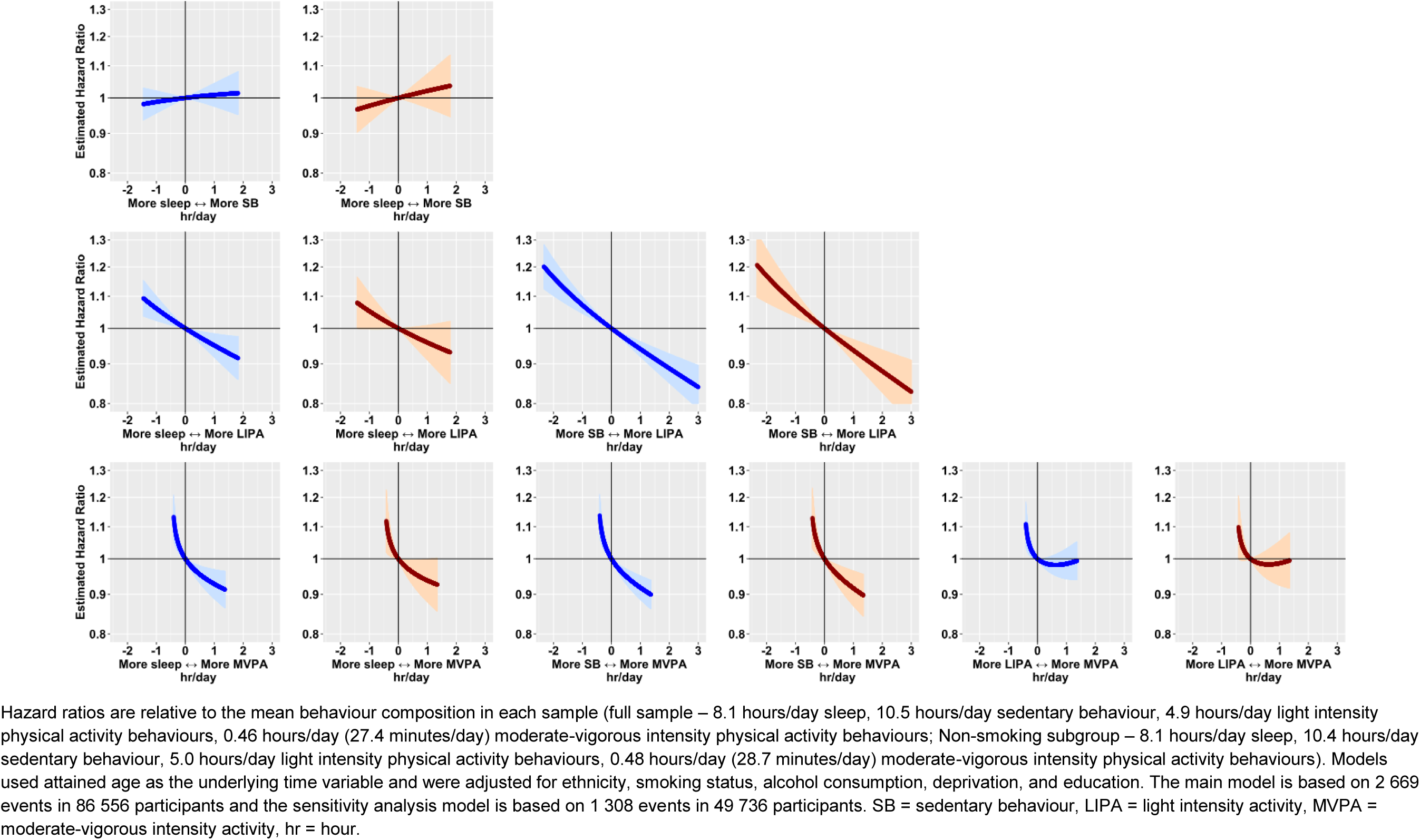
Hazard ratios for all behaviour pairs and incident physical-activity-related cancer risk for estimated using a multivariable-adjusted Cox regression model in 86 556 UK Biobank participants before (blue) and after restricting to never smokers (red, N=49 736).

**eFigure 7.**
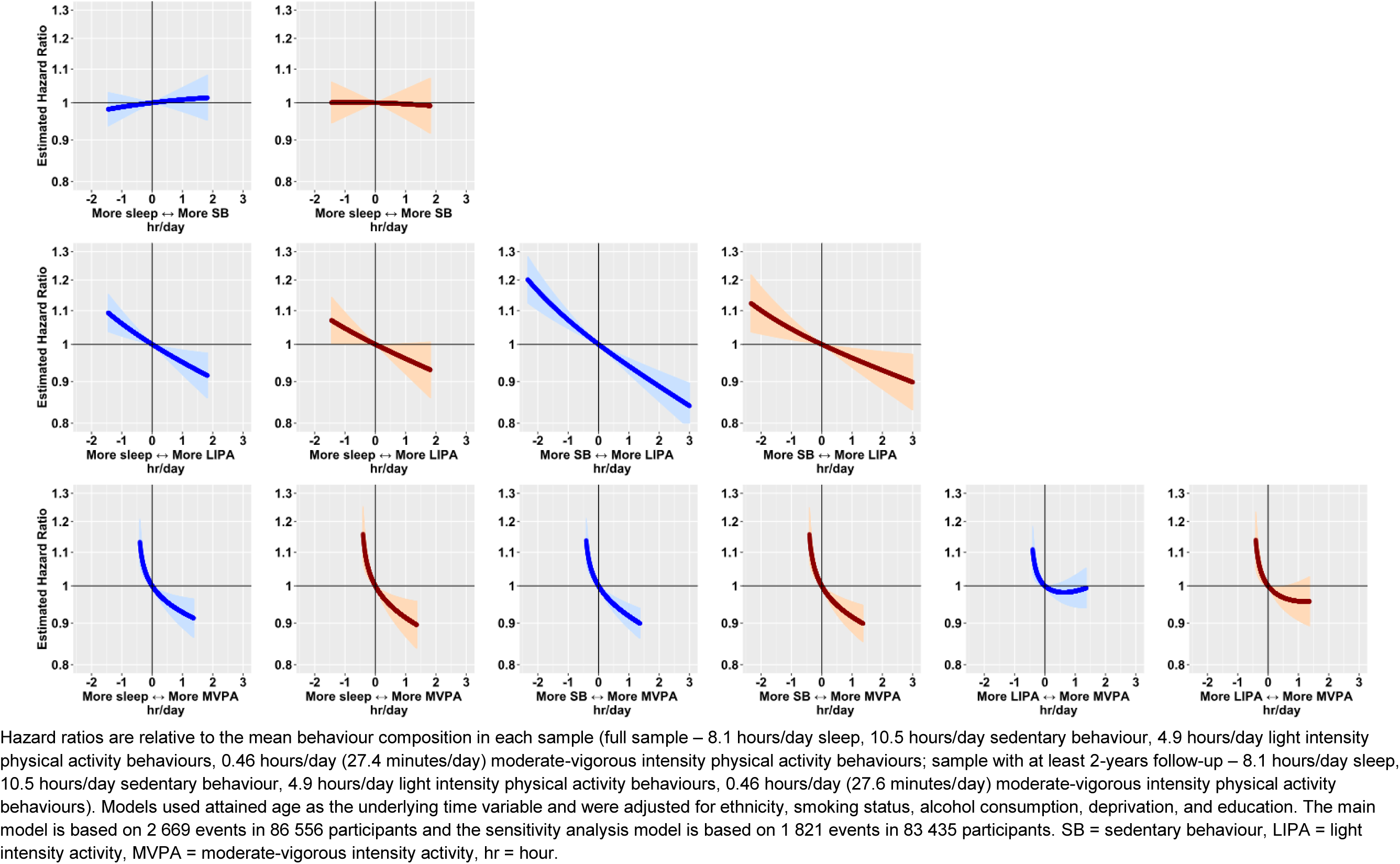
Hazard ratios for all behaviour pairs and incident physical-activity-related cancer risk for estimated using a multivariable-adjusted Cox regression model in 86 556 UK Biobank participants before (blue) and after removing the first two years of follow-up (red, N=83 435).

## Notes

### Funding Statement

Shreves Saint-Maurice Moore and Matthews were supported by the National Institutes of Health Intramural Research Program
Shreves is also supported by the National Institutes of Health Oxford Cambridge Scholars Program
Walmsley is supported by HDR UK an initiative funded by UK Research and Innovation Department of Health and Social Care (England) and the devolved administrations
Papier is supported by Cancer Research UK (grant number C16077/A29186)
Chan is supported by Novo Nordisk Doherty and Small are supported by the Wellcome Trust (223100/Z/21/Z)
Doherty is also supported by Novo Nordisk Swiss Re and the British Heart Foundation Centre of Research Excellence (grant number RE/18/3/34214)
Travis is supported by Cancer Research UK (grant number C8221/A29017)
The funders/sponsors had no role in the design of this study or the analysis and interpretation of the results or drafting of the manuscript

### Author Declarations

UK Biobank particiapnt provided written informed consent. The study was approved by the National Information Governance Board for Health and Social Care and the National Health Service North West Multicentre Research Ethics Committee (06/MRE08/65).

